# Mathematical modeling of the transmission of SARS-CoV-2 – Evaluating the impact of isolation in São Paulo State (Brazil) and lockdown in Spain associated with protective measures on the epidemic of covid-19

**DOI:** 10.1101/2020.07.30.20165191

**Authors:** Hyun Mo Yang, Luis Pedro Lombardi Junior, Fabio Fernandes Morato Castro, Ariana Campos Yang

## Abstract

Coronavirus disease 2019 (covid-19), with the fatality rate in elder (60 years old or more) being much higher than young (60 years old or less) patients, was declared a pandemic by the World Health Organization on March 11, 2020. Taking into account this age-dependent fatality rate, a mathematical model considering young and elder subpopulations was formulated based on the natural history of covid-19 to study the transmission of the SARS-CoV-2. This model can be applied to study the epidemiological scenario resulting from the adoption of isolation or lockdown in many countries to control the rapid propagation of covid-19. We chose as examples the isolation adopted in São Paulo State (Brazil) in the early phase but not at the beginning of the epidemic, and the lockdown implemented in Spain when the number of severe covid-19 cases was increasing rapidly. Based on the data collected from São Paulo State and Spain, the model parameters were evaluated and we obtained higher estimation for the basic reproduction number *R*_0_ (9.24 for São Paulo State, and 8 for Spain) compared to the currently accepted estimation of *R*_0_ around 3. The model allowed to explain the flattening of the epidemic curves by isolation in São Paulo State and lockdown in Spain when associated with the protective measures (face mask and social distancing) adopted by the population. However, a simplified mathematical model providing lower estimation for *R*_0_ did not explain the flattening of the epidemic curves. The implementation of the isolation in São Paulo State before the rapidly increasing phase of the epidemic enlarged the period of the first wave of the epidemic and delayed its peak, which are the desirable results of isolation to avoid the overloading in the health care system.

## Introduction

Coronavirus disease 2019 (covid-19) is caused by severe acute respiratory syndrome coronavirus 2 (SARS-CoV-2), which is a strain of the RNA-based SARS-CoV-1. This SARS-CoV-2 (new coronavirus) can be transmitted by droplets that escape the lungs through coughing or sneezing and infect humans (direct transmission), or they are deposited in surfaces and infect humans when in contact with this contaminated surface (indirect transmission) [1] [2]. These routes of transmission resulted in a rapid spreading of this virus, and the World Health Organization (WHO) declared covid-19 a pandemic on March 11, 2020.

This virus enters into susceptible persons through the nose, mouth, or eyes, and infects cells in the respiratory tract. Covid-19 in mild form presents fever, dry cough, chills, malaise, muscle pain, and sore throat, in moderate form presents fever, respiratory symptoms, and radiographic characteristics, and in severe form manifests dyspnea, low oxygen saturation, and may evolve to respiratory failure, and multiple organ failure. In general, the fatality rate in elder patients (60 years or more) is much higher than those with 60 years or less [3].

Currently, there is not a vaccine, neither an effective treatment. Hence, at the beginning of covid-19 outbreaks, isolation was the main way of controlling the dissemination of the new coronavirus in a population [4]. However, there is evidence that individual (protection of mouth and nose using face mask and protection of eyes, and washing hands with alcohol and gel) and collective (social distancing) protective measures diminish the transmission of covid-19 [5]. The decrease in the incidence of covid-19 by isolation, known as flattening the curve of an epidemic, can be quantified by mathematical modeling.

Initially, computational models (especially, agent-based model) to describe influenza epidemics were adapted and applied to estimate the spreading of SARS-CoV-2. Koo *et al*. [6] used such a model to study the propagation of the new coronavirus in Singapore, assuming that the basic reproduction number, denoted by *R*_0_, was around 2. The same approach was done by Ferguson *et al*. [7] to investigate the impact of non-pharmaceutical interventions (isolation of susceptible persons) named mitigation and suppression. Briefly, mitigation reduces the basic reproduction number *R*_0_, but not lower than one, while suppression reduces the basic reproduction number lower than one. They simulated their model assuming *R*_0_ around 2.5, and predicted the numbers of severe cases and deaths due to covid-19 without interventions and compared them with those numbers when isolation (mitigation or suppression) was implemented in a population. However, instead of assuming a certain value for *R*_0_, Li *et al*. [8] performed a stochastic simulation of SEIR (susceptible-exposed-infectious-recovered) model incorporating the rapid dissemination of new coronavirus due to undocumented infections, and estimated the effective reproduction number *R*_*ef*_ around 2.4. The basic and effective reproduction numbers *R*_0_ and *R*_*ef*_ must be retrieved from the model, and they are defined in Materials and Methods.

The herd immunity jumps down *R*_*ef*_ by the vaccination [9], as the isolation of the susceptible persons (non-pharmaceutical interventions [7]) jumps it down. Hence, we name as “herd protection” the protection conferred to susceptible persons by isolation, and the adoption of individual (face mask and constant hygiene by washing hands with alcohol and gel) and collective (social distancing) protective measures by the non-isolated (circulating) population.

Mathematical models based on a well documented natural history of the disease allow us to understand the progression of viral infection and provide mathematical expression to estimate *R*_0_, which is related to the magnitude of efforts to eradicate an infection [9]. When a simple SIR model is considered to describe the covid-19 epidemic, it is expected to estimate *R*_0_ around 3. Fortunately, the knowledge about the natural history of covid-19 is being improved every day as the epidemic evolves, and, as a consequence, mathematical modeling can be benefited by incorporating novel aspects of this epidemic. In [10], a mathematical model encompassing two subpopulations based on the different fatality rates in young (60 years old or less) and elder (60 years old or more) subpopulations was developed aiming to study the impacts of the isolation and further release on the epidemic of covid-19. That model was applied to São Paulo State (Brazil) to describe the epidemiological scenario considering intermittent pulses in isolation and release. We improved that model allowing the transmission of the infection by persons presenting mild covid-19 symptoms, and incorporating the protective measures that reduce the transmission of the virus.

The model proposed here is applied to evaluate the impacts of herd protection on the epidemics in São Paulo State and Spain. The widespread of covid-19 in Spain led to the adoption of lockdown, which is an extreme measure to control the quick increase of an epidemic. São Paulo State, however, implemented the isolation in the population to avoid critical epidemiological scenarios occurring in Spain and Italy. Based on the data collection of severe covid-19 cases and deaths from São Paulo State and Spain, we estimate the model parameters, the proportion of the population in isolation/lockdown, and the reduction in the transmission rates by the adoption of the protective measures. The relatively early adoption of the isolation in São Paulo State resulted in the enlarging of the period of the first wave of the epidemic and delaying its peak. Based on the description of the epidemic under herd protection, the model can provide epidemiological scenarios of the strategies of release that can help guide public health policies by decision-makers.

## Results and Discussion

In Materials and Methods, we present a mathematical model to describe the transmission of SARS-CoV-2, the description of collections of data from São Paulo State and Spain, and the values assigned to the model parameters. This model is applied to describe the epidemiological scenarios of isolation in São Paulo State and lockdown in Spain, and we obtained the basic reproduction number *R*_0_ and also the effective reproduction number *R*_*ef*_.

From Materials and Methods, we observed three distinct trends in the accumulated covid-19 data. These three periods in São Paulo State describe the epidemic occurring naturally, with isolation, and isolation with protective measures (herd protection). In Spain, the three periods represent the natural epidemic, the epidemic during the transition from natural to lockdown, and the epidemic occurring in lockdown. Taking into account these three trends shown by the covid-19 data, we evaluate the model parameters *β*_*y*_, *β*_*o*_, *k, ε*, and *ω*. However, the additional mortality rates *α*_*y*_ and *α*_*o*_ are evaluated considering all three periods. We then assess the effects of herd protection (non-pharmaceutical interventions) on the flattening of epidemic curves in São Paulo State and Spain.

The first period of data corresponding to the natural epidemics of covid-19 in São Paulo State and Spain is a unique opportunity to estimate the basic reproduction number *R*_0_. Based on this estimation, all subsequent interventions applied to control epidemics can be explained. Indeed, the magnitude of the jump down in the effective reproduction number *R*_*ef*_ is proportional to the control intervention. The similitude between mass vaccination and isolation is the proportion of susceptible individuals being protected from infection, which results in the jump down of *R*_*ef*_.

### Covid-19 in São Paulo State – Isolation

São Paulo State has 44.6 million inhabitants with 15.3% of elder population (60 years old or more) [11], and the demographic density is 177*/km*^2^ [12]. The first confirmed case of covid-19 occurred on February 26, the first death due to covid-19 on March 16, and on March 24, São Paulo State implemented the isolation of people in non-essential activities. The lockdown implemented in Spain and Italy due to their critical epidemiological scenarios led to the adoption of isolation in São Paulo State. We evaluate the model parameters using daily collected data (see Figures 17 and 18), and describe the epidemiological scenario with isolation.

#### Parameters evaluation

Using data collected in São Paulo State from February 26 to May 7, we evaluate the transmission rates (*β*_*y*_ and *β*_*o*_), the proportion in the isolated population (*k*), reduction in the transmission rates due to the protective measures adopted by the circulating population (*ε*), and the additional mortality rates (*α*_*y*_ and *α*_*o*_).

#### Natural epidemic – Evaluating the transmission rates

The effects of isolation implemented on March 24 are expected to appear later on the daily registered cases of severe covid-19 (the sum of incubation and pre-diseased infection periods (see Table 2) is 9.8 days). Hence, the data from February 26 to April 3 of severe covid-19 cases portray the natural epidemic, that is, the transmission of infection is occurring without any kind of intervention. This epidemiological scenario fulfills the definition of the basic reproduction number *R*_0_ (entire population is susceptible in the absence of constraints), allowing its estimation.

We evaluate the transmission rates taking into account the confirmed cases from February 26 (*t*_1_) to April 3 (*t*_38_), and using equation (33). The evaluated values are *β*_*y*_ = 0.78 and *β*_*o*_ = 0.90 (both in *days*^*−*1^), where *ψ* = 1.15, resulting in the basic reproduction number *R*_0_ = 9.24 (partials *R*_0*y*_ = 7.73 and *R*_0*o*_ = 1.51), according to equation (30).

Figure 1(a) shows the estimated curve of Ω and the observed data, plus two curves with lower transmission rates: *β*_*y*_ = 0.59 and *β*_*o*_ = 0.68 (both in *days*^*−*1^), with *R*_0_ = 6.99 (partials *R*_0*y*_ = 5.84 and *R*_0*o*_ = 1.16); and *β*_*y*_ = 0.43 and *β*_*o*_ = 0.50 (both in *days*^*−*1^), with *R*_0_ = 5.09 (partials *R*_0*y*_ = 4.26 and *R*_0*o*_ = 0.84). Figure 1(b) shows the extended curves of Ω, from equation (14), which approach asymptotes (or plateaus) indicating the end of the first wave of the epidemic. For *R*_0_ = 9.24, 6.99, and 5.09, the curves Ω reach values with little difference on September 13, respectively, 946, 400, 945, 700, and 941, 500. For *R*_0_ = 9.24, the curves for young (Ω_*y*_) and elder (Ω_*o*_) persons approach on September 13 values, respectively, 605, 300 and 341, 100.

**Figure 1:**
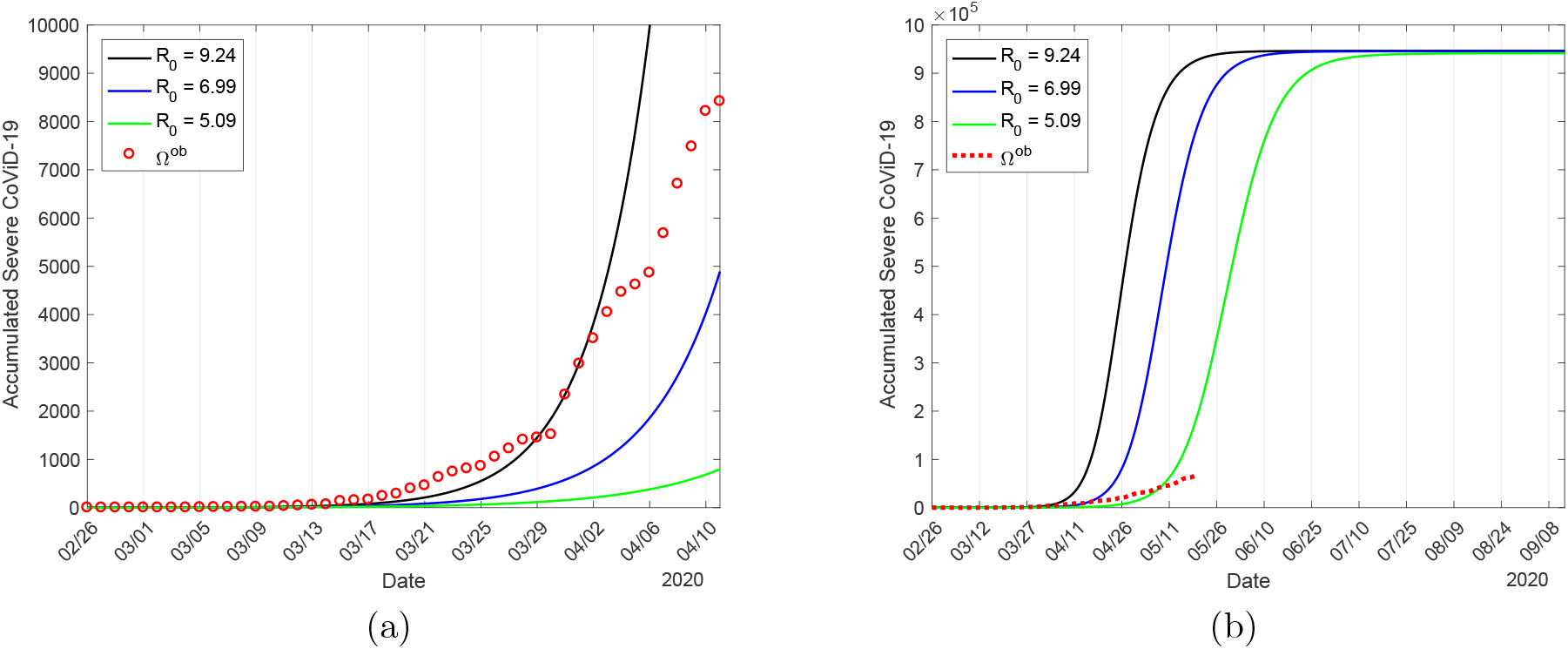
The estimated curve of the accumulated number of severe covid-19 cases Ω in natural epidemic and observed data in São Paulo State, plus two curves with lower transmission rates: *β*_*y*_ = 0.59 and *β*_*o*_ = 0.68 (*days*^*−*1^), with *R*_0_ = 6.99, and *β*_*y*_ = 0.43 and *β*_*o*_ = 0.50 (*days*^*−*1^), with *R*_0_ = 5.09 (a), and extended curves of Ω (b).

We stress the fact that, if the observed data are fitted without caution about interventions, that is, using all data indistinctly, someone could estimate the basic reproduction number to be *R*_0_ = 5.09 or less (notice that near the horizontal axis of Figure 1(b), the observed data do not approach the curve of *R*_0_ = 5.09, rather lower *R*_0_).

It is accepted that SARS-CoV-2 is an airborne infection [2]. For instance, for the rubella infection, using seroprevalence data [13] and dealing with the SEIR model in the steady-state, the estimation was *R*_0_ = 6.71 [14]. That estimation was done before the mass vaccination against rubella infection, which was the reason to assume that the estimation was the basic reproduction number. However, the rubella virus was circulating probably not in a steady-state, then the estimation would not be *R*_0_, but the effective reproduction number *R*_*ef*_, given by equation (29), and the actual value of *R*_0_ must be higher than 6.71. Moreover, Caieiras City in São Paulo State had 30, 000 inhabitants with a demographic density of 264*/km*^2^ in 1990, which indicates that *R*_0_ = 6.71 is under-estimation for rubella transmission in São Paulo State. As we pointed out in Materials and Methods, we have only two moments during the epidemic when *R*_0_ can be estimated from data: At the beginning without any kind of constraints (during the natural epidemic), and when the steady-state is reached, which in general occurs after a long time [14].

In Discussion, we present more arguments to demonstrate that the currently accepted lower *R*_0_ (around 3) does not explain the covid-19 epidemic.

#### Epidemic with isolation – Evaluating the proportion of the population in isolation

Isolation was introduced on March 24 (*t*_1_), and we evaluate the proportion in isolation taking into account the confirmed cases of covid-19 until April 12 (*t*_20_), and using equation (33).

To evaluate the proportion of the population in isolation, we fix the transmission rates *β*_*y*_ = 0.78 and *β*_*o*_ = 0.90 (both in *days*^*−*1^), and vary *k* = 0, 0.4, 0.53, 0.6, 0.7, and 0.8, where *k*_*mean*_ = 0.53 is the average proportion of persons in isolation from March 24 to May 3 (see Figure 18). We observe that *k* = 0.4 and 0.6 fit part of the observed data, while *k* = 0.7 does not. Hence, we chose *k* = *k*_*mean*_ = 0.53 as the value that explains the isolation in São Paulo State. Figure 2 shows the curves of Ω for different proportions in isolation in São Paulo State and the observed data (a), and the extended curves of Ω (b).

**Figure 2:**
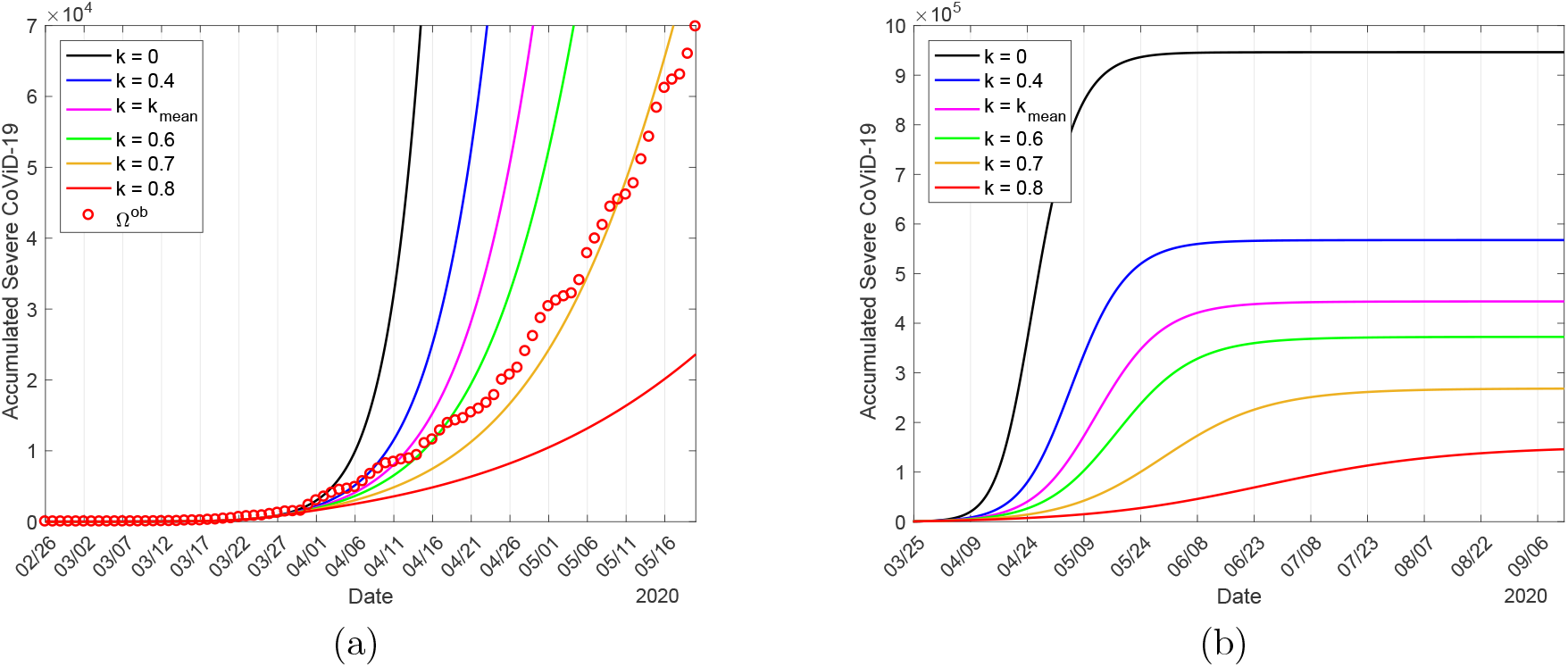
The curves of Ω for the proportions in isolation in São Paulo State *k* = 0, 0.4, 0.53, 0.6, 0.7, and 0.8, and observed data (a), and the extended curves of Ω (b).

From Figure 2(b), the curves Ω approach plateau, and the values on September 13 for *k* = 0, 0.4, 0.53, 0.6, 0.7, and 0.8 are, respectively, 964, 600, 567, 600 (60%), 444, 000 (47%), 372, 500 (40%), 268, 200 (28%), and 146, 000 (15%). The percentage between parentheses is the ratio Ω(*k*)*/*Ω(0). For *k* = *k*_*mean*_, the values for Ω_*y*_ and Ω_*o*_ are, respectively, 283, 600 and 160, 400.

As we have pointed out in the description of the data (Materials and Methods), the observed proportion in isolation delayed by approximately 9 days indeed affected the daily incidence of covid-19. Moreover, the chosen proportion *k*_*mean*_ is the average proportion of isolation in São Paulo State. The number of accumulated cases Ω decreases as *k* increases, showing that the isolation of the population decreases the transmission of covid-19, flattening the epidemic curve. This decrease can be assessed also by equation (31), which is the reduction in the effective reproduction number by isolation *R*_*r*_.

When isolation is implemented during the epidemic, those who are harboring the virus can be found in the isolated population. At the time of the beginning of isolation in São Paulo State on March 24, we have

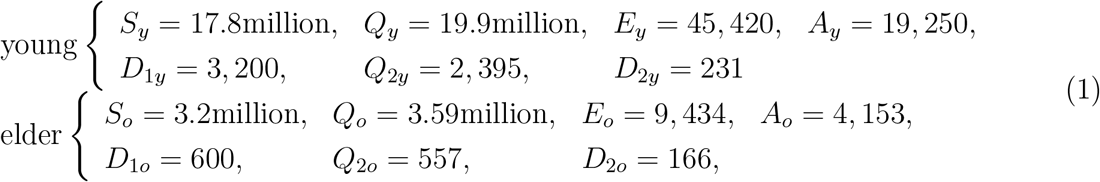

with *Q*_1*y*_ = *Q*_1*o*_ = *Q*_3*y*_ = *Q*_3*o*_ = 0 and *I* = 10, 032. When 53% (*k* = 0.53) of the infectious persons in each class is transferred to the isolated classes, the total number of isolated persons harboring virus 43, 490 can trigger a new epidemic in the population in isolation. However, if the transmission rate is low in the isolated population [15], the number of severe covid-19 cases is around 1% compared with the peak 67, 140 shown below. Hence, we assume that SARS-CoV-2 is not transmitting in the isolated population, and we do not evaluate *ω*.

#### Epidemic with isolation and protective measures – Evaluating the reduction in the transmission rates

On April 13, 20 days after the beginning of isolation, we observe the first point leaving completely the curve. This new trend can not be explained by an increased proportion of isolation (see Figure 18). To take into account this new tendency of data, in [10] we hypothesized that the using of face mask (protection of mouth and nose), protection of eyes, constant hygiene (washing hands with alcohol and gel) and social distancing may decrease the transmission of infection [5] [16]. Differently to isolation, these protective measures reduce the transmission rates as can be seen in equation (3).

We consider that on April 4, 9 days before the observed points leaving consistently the estimated curve of the epidemic with isolation, the protective measures were adopted by population, which reduced the transmission rates from *β*_*y*_ and *β*_*o*_ to 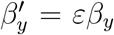 and 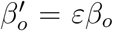. We fix the transmission rates *β*_*y*_ = 0.78 and *β*_*o*_ = 0.90 (both in *days*^*−*1^), and the proportion in isolation *k* = 0.53 to evaluate the protective factor *ε* taking into account data from April 4 (*t*_1_) to May 7 (*t*_34_), and using equation (33).

We vary *ε* = 0.8, 0.7, 0.6, 0.5, and 0.4, and the value *ε* = 0.5 is chosen to represent the protective measures adopted in São Paulo State. Figure 3 shows the curves of Ω for *ε* = 1, 0.8, 0.7, 0.6, 0.5, and 0.4 plus natural epidemic (*k* = 0) and the observed data (a), and the extended curves of Ω for *ε* = 1, 0.7, 0.6, 0.5, and 0.4 (b). The curves Ω approach plateau, and on November 2, the values for *ε* = 1, 0.7, 0.6, 0.5 and 0.4 are, respectively, 444, 000, 427, 900 (97%), 413, 500 (93%), 386, 600 (87%), and 331, 200 (75%). The percentage between parentheses is the ratio Ω(*ε*)*/*Ω(0). For *ε* = 0.5, the values for Ω_*y*_ and Ω_*o*_ are, respectively, 243, 800 and 142, 800, which are decreased by 86% and 89% in comparison with isolation alone as intervention.

**Figure 3:**
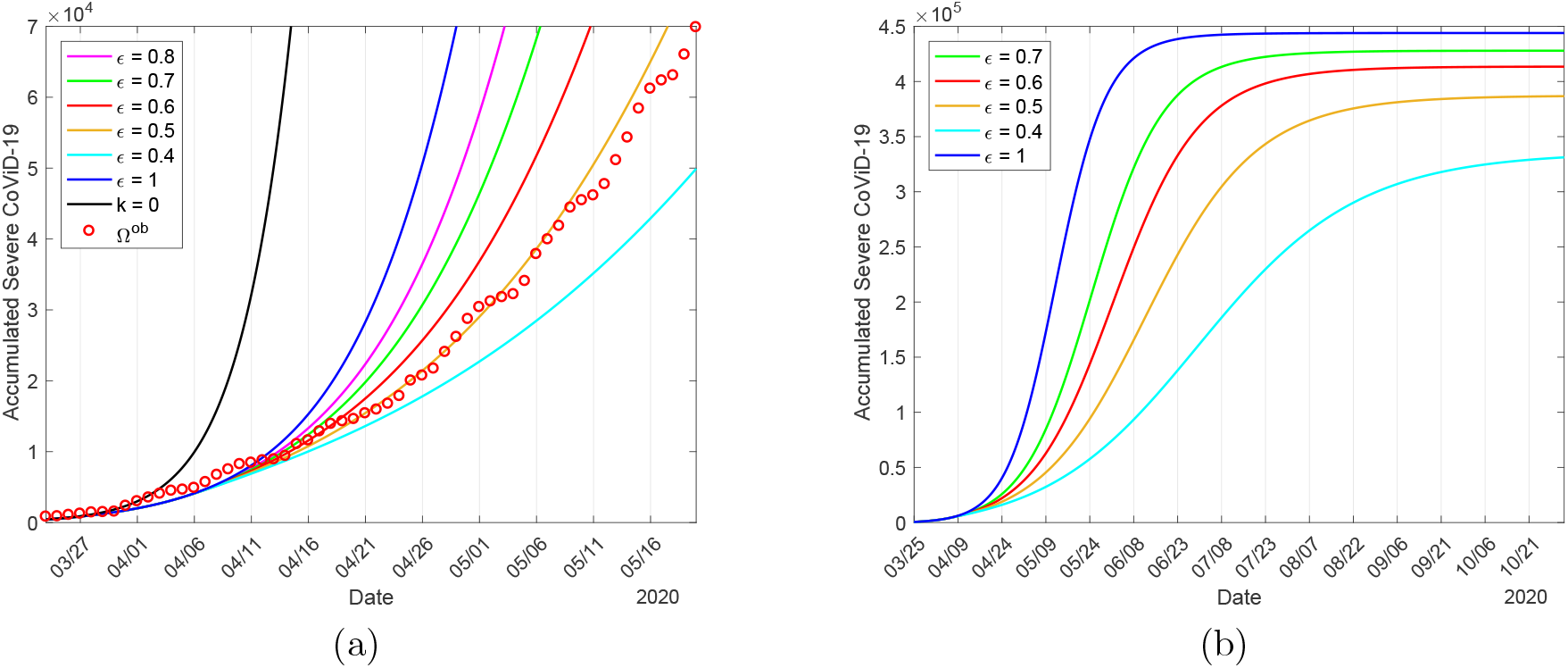
The curves of Ω for the protective measures *ε* = 1, 0.8, 0.7, 0.6, 0.5 and 0.4, plus natural epidemic (*k* = 0) and the observed data (a), and the extended curves of Ω for *ε* = 1, 0.7, 0.6, 0.5 and 0.4 (b).

The number of accumulated cases Ω decreases as *ε* decreases (1*−ε* is the effectiveness of protective measures), showing that the protective measures adopted by the population decreased the transmission of covid-19, flattening the epidemic curve. This decrease can be assessed also by equation (32), which is the reduction in the effective reproduction number by protective measures *R*_*p*_.

#### Evaluating the additional mortality rates

We estimate the additional mortality rates *α*_*y*_ = Γ*α*_0_ and *α*_*o*_ taking into account confirmed deaths from March 16 (*t*_1_) to May 20 (*t*_66_), and using equation (34). We fix the previously estimated transmission rates *β*_*y*_ = 0.78 and *β*_*o*_ = 0.90 (both in *days*^*−*1^), the proportion in isolated population *k* = 0.53, and the protective factor *ε* = 0.5, to evaluate *α*_0_. As we pointed out in Materials and Methods, we fix Δ = 15 *days* and let Γ = 0.26 in São Paulo State (74% of deaths are occurring in elder persons with severe covid-19 [17]). The evaluated additional mortality rates are *α*_*y*_ = 0.00185 and *α*_*o*_ = 0.0071 (both in *days*^*−*1^).

Figure 4 shows the estimated curve of Π, from equation (16), and the observed death data (a), and the extended curves of the number of covid-19 deaths for young Π_*y*_, elder Π_*o*_, and total Π = Π_*y*_ + Π_*o*_ persons (b). The estimated curves Π_*y*_, Π_*o*_, and Π reach plateaus, and on November 2 the values are, respectively, 5, 280 (2.2%), 18, 500 (13%), and 23, 780 (6.0%). The percentage between parentheses is the severe covid-19 case fatality rate Π*/*Ω, Ω being given in Figure 3(b).

**Figure 4:**
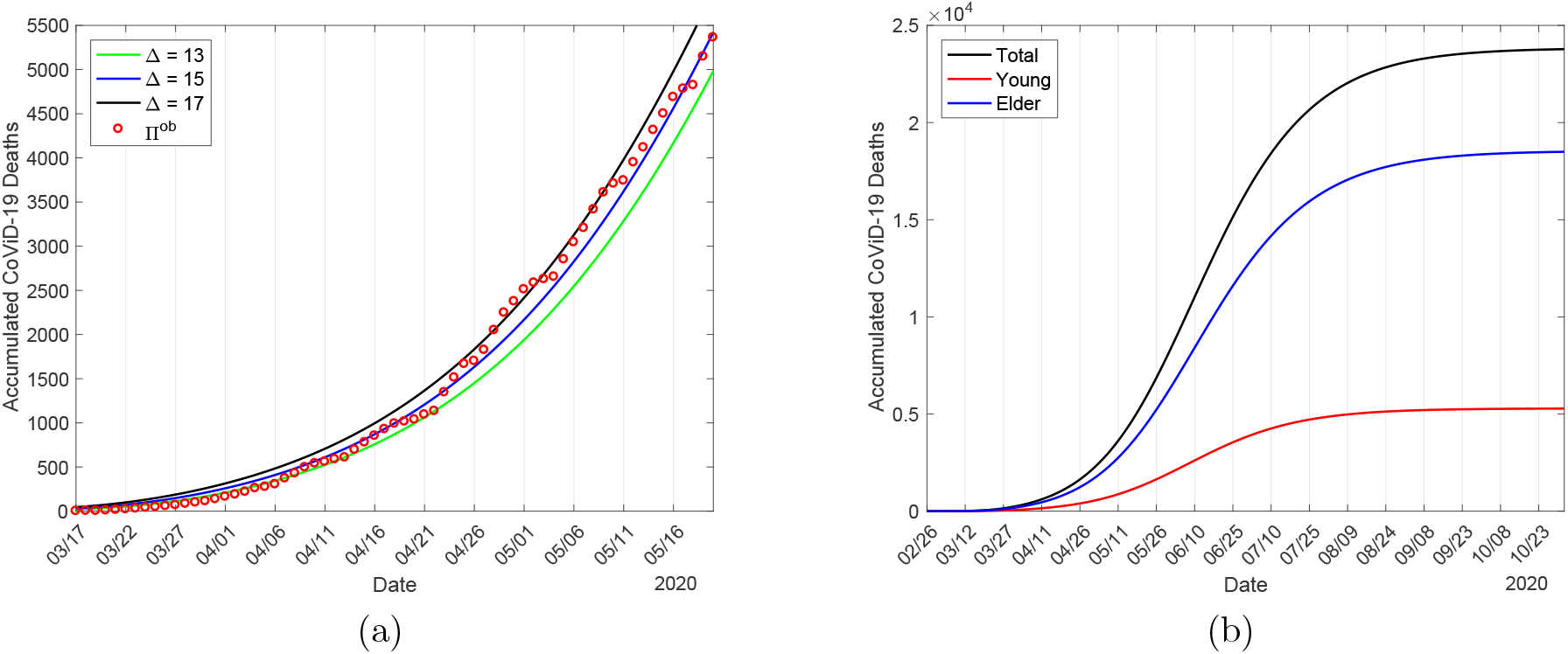
The estimated curves of the accumulated deaths due to covid-19 Π for Δ = 13, 15 and 17 *days*, and the observed data in São Paulo State (a), and the extended curves for young Π_*y*_, elder Π_*o*_, and total Π = Π_*y*_ + Π_*o*_ persons for Δ = 15 *days* (b).

At the end of the first wave of the epidemic, 78% of all deaths occurred in elder subpopulation. The number of the new cases of covid-19, from equation (13), for young Φ_*y*_, elder Φ_*o*_, and total Φ = Φ_*y*_ + Φ_*o*_ persons are, respectively, 15.3 million, 2.86 million and 18.16 million. The infection fatality rate (Π*/*Φ) in young, elder and all persons are, respectively, 0.031%, 0.65%, and 0.13%.

#### Epidemiological scenario with isolation

We present the epidemiological scenario with isolation using all previously estimated parameters: The transmission rates *β*_*y*_ = 0.78 and *β*_*o*_ = 0.90 (both in *days*^*−*1^), giving *R*_0_ = 9.24; the additional mortality rates *α*_*y*_ = 0.00185 and *α*_*o*_ = 0.0071 (both in *days*^*−*1^); the proportion in isolation of susceptible persons *k* = 0.53; and the protective factor *ε* = 0.5 reducing the transmission rates to 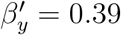 and 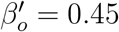 (both in *days*^*−*1^), giving *R*_0_ = 4.62.

In Figure 5, we show the effects of interventions on the dynamics of the new coronavirus. As interventions are added (isolation followed by protective measures), we observe decreasing in the peaks of severe covid-19 *D*_2_, which move to the right. Figure 5(a) shows the curves of the natural epidemic, epidemic considering only isolation, and epidemic occurring with isolation and protective measures. In Figure 5(b), we show the number of immune persons *I* corresponding to the three cases shown in Figure 5(a). The curves have sigmoid-shape.

**Figure 5:**
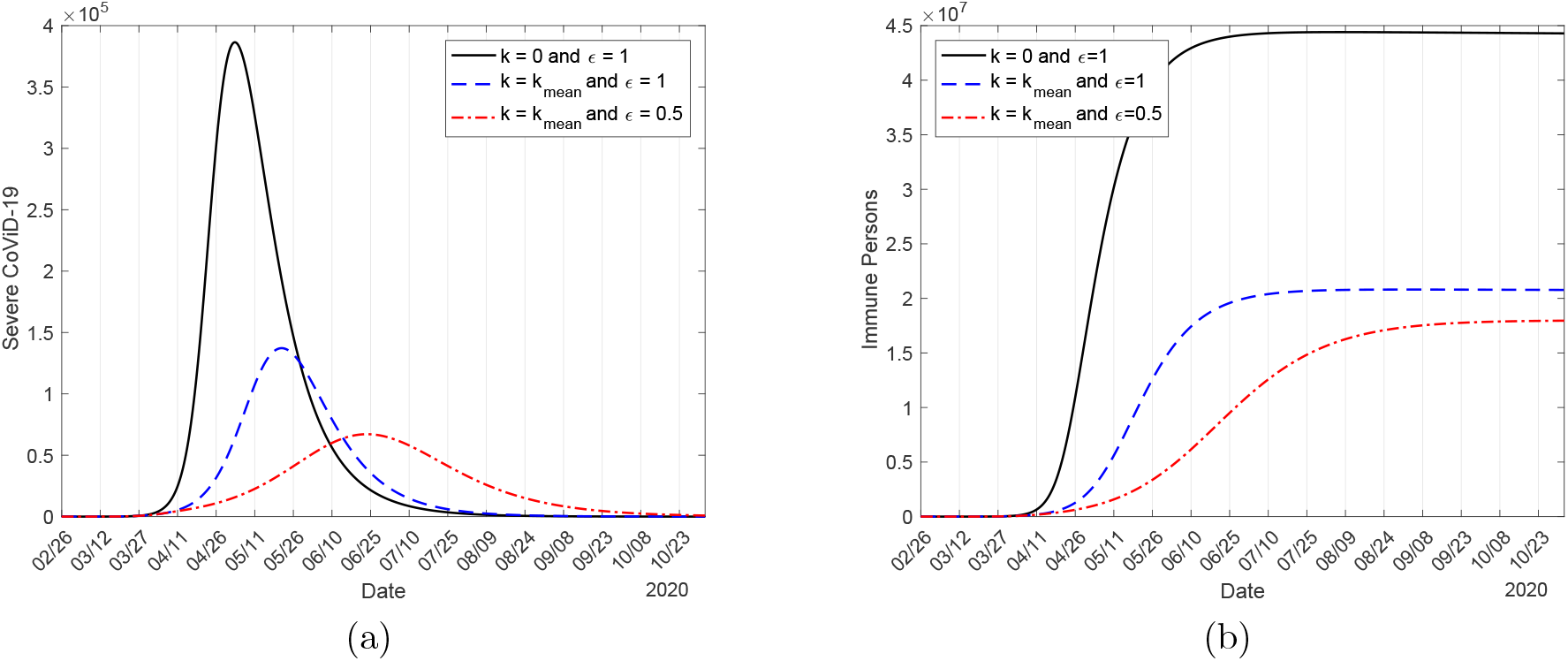
The curves of the natural epidemic (*k* = 0 and *ε* = 1), epidemic considering only isolation (*k* = 0.53 and *ε* = 1), and epidemic occurring with isolation and protective measures (*k* = 0.53 and *ε* = 0.6) (a), and the number of immune persons *I* (b).

In the absence of interventions (*k* = 0 and *ε* = 1), on June 15, the numbers of immune persons *I*_*y*_, *I*_*o*_, and *I* increase from zero to, respectively, 36.92 million, 6.505 million and 43.43 million. When interventions (isolation and protective measures) are adopted, the numbers are 6.12 million (16.6%), 1.14 million (16.5%), and 7.26 million (16.7%) on June 15. The percentage between parentheses is the ratio between with and without interventions *I*(*k, ε*)*/I*(0, 1).

Let us compare the peak of *D*_2_. The peaks for young, elder and total persons in the natural epidemic are, respectively, 224, 200, 162, 200, and 386, 400, occurring on May 2, 4, and 3. Considering isolation alone (*k* = 0, 53 and *ε* = 1), the peaks for young, elder and total persons are, respectively, 77, 320 (34%), 60, 020 (37%), and 137, 200 (36%), which occur on May 21, 22, and 21. Considering isolation and protective measures (*k* = 0, 53 and *ε* = 0.5), the peaks for young, elder and total persons are, respectively, 36, 010 (16%), 31, 160 (19%), and 67, 140 (18%), which occur on June 22, 24, and 23. The percentage between parentheses is the ratio between with and without interventions *D*_2_(*k, ε*)*/D*_2_(0, 1).

Due to isolation and protective measures, so many people remain as susceptible. In Figure 6 we show circulating susceptible persons *S*_*y*_, *S*_*o*_ and *S* = *S*_*y*_ + *S*_*o*_ (a), and circulating plus isolated susceptible persons 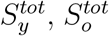 and 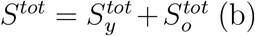, using equation (12). Remember that 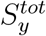 differs from *S*_*y*_ just after the introduction of isolation (March 24).

**Figure 6:**
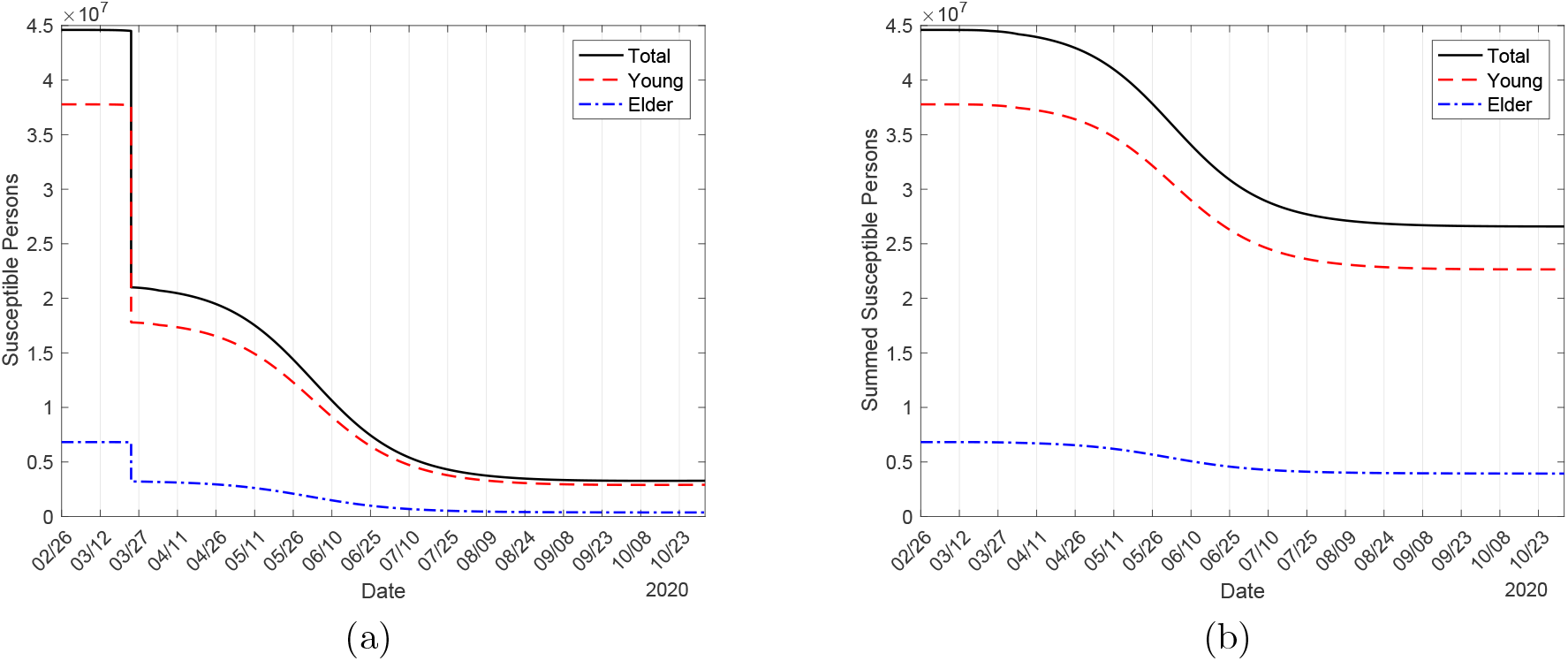
The circulating susceptible young *S*_*y*_, elder *S*_*o*_ and total *S* = *S*_*y*_ + *S*_*o*_ persons (a), and the sum of the circulating and isolated susceptible populations 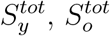 and 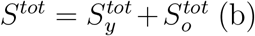.

At the end of the first wave of the epidemic, the numbers of susceptible persons *S*_*y*_, *S*_*o*_, and *S* = *S*_*y*_ + *S*_*o*_ without any interventions (*k* = 0 and *ε* = 1) are, respectively, 37, 000, 210 and 37, 210, and the numbers of susceptible persons with interventions (isolation and protective measures) are 8.15 million (22, 027%), 1.3 million (619, 047%), and 9.5 million (25, 531%), for young, elder and total persons, respectively. For 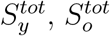 and 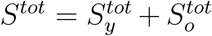, we have, respectively, 28 million (75, 676%), 4.9 million (2, 333, 333%), and 33 million (88, 686%). The percentage between parentheses is the ratio between with and without interventions *S*(*k, ε*)*/S*(0, 1). Observe that, on June 15, the sum of the susceptible persons in circulation and those in isolation is such that there are more than 750-time and 23, 000-time, respectively, susceptible young and elder persons in comparison with epidemic without any intervention. Hence, if all persons are released without planning, the second wave will be intense, infecting much more elder persons. In the absence of vaccine and effective treatment, interventions to reduce the transmission of SARS-CoV-2 must be continued for a long time to avoid the rebounding of the epidemic or a second wave.

It is important to estimate the basic reproduction number, which portrays the beginning and ending phases of an epidemic [18]. During the epidemic, however, the effective reproduction number determines the intensity of transmission of the infection. We use the approximate effective reproduction number *R*_*ef*_, given by equation (29), to follow the trend of the epidemic, remembering that *R*_*ef*_ *>* 1 implies epidemic in expansion, while *R*_*ef*_ *<* 1, in contraction. Figure 7 illustrates the effective reproduction number *R*_*ef*_ and *D*_2_ during the epidemic, with (a) and without (b) intervention. To be fitted together in the same frame with *R*_*ef*_, the curve of *D*_2_ is divided by 7, 000 (a) and 40, 000 (b). The curve of *R*_*ef*_ follows the shape of susceptible persons as shown in Figure 6, as expected. At the peak of the epidemic, the effective reproduction number is lower than one, hence we have *R*_*ef*_ = 1 occurring on June 14 (a) and April 6 (b).

**Figure 7:**
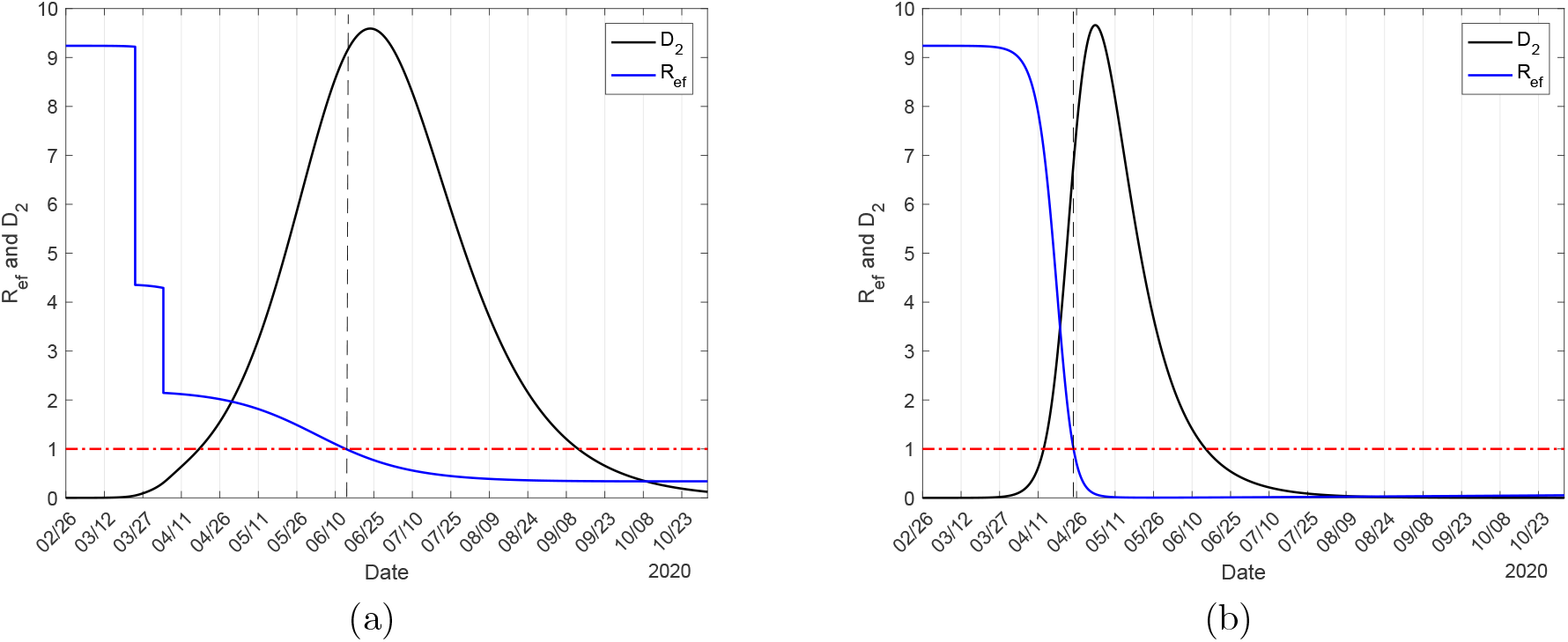
The effective reproduction number *R*_*ef*_ for epidemic with isolation and protective measures (a), and natural epidemic (b) in São Paulo State. The number of severe covid-19 cases *D*_2_ must be multiplied by 7, 000 (a) and 40, 000 (b).

As the epidemic evolves, the effective reproduction number varies as shown in Figure 7(a). At the beginning of the epidemic, on February 26, we have *R*_*ef*_ = *R*_0_ = 9.24, on March 24, a jump down occurred to *R*_*ef*_ = 4.35 due to the isolation, and a new jump down occurs to *R*_*ef*_ = 2.15 on April 4 when protective measures were adopted. On June 15, when the release will begin, we have *R*_*ef*_ = 0.98, but in the ascending phase of the epidemic. The knowledge of *R*_*ef*_ could help public health authorities to plan the strategies of release.

We used the accumulated data shown in Figure 17(b) and Ω given by equation (14) to estimate the transmission rates *β*_*y*_ and *β*_*o*_, the proportion in isolation *k*, and the protective factor *ε*. The curve labeled *ε* = 0.5 in Figure 3(b) is the estimated curve Ω, from which the curve of severe cases *D*_2_ was derived, corresponding to the most flattened curve shown in Figure 5(a). Now, from the estimated curve of Ω, we derive the daily cases Ω_*d*_ given by equation (15). In Figure 8(a), we show the calculated curve Ω_*d*_ and daily cases presented in Figure 17(a). In Figure 8(b), we show the initial part of the estimated curve Ω with observed data Ω^*ob*^, the extended Ω_*d*_ and daily observed cases 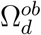, and severe cases *D*_2_. The peaks of Ω_*d*_ and *D*_2_ occur, respectively, on June 12 and 23.

**Figure 8:**
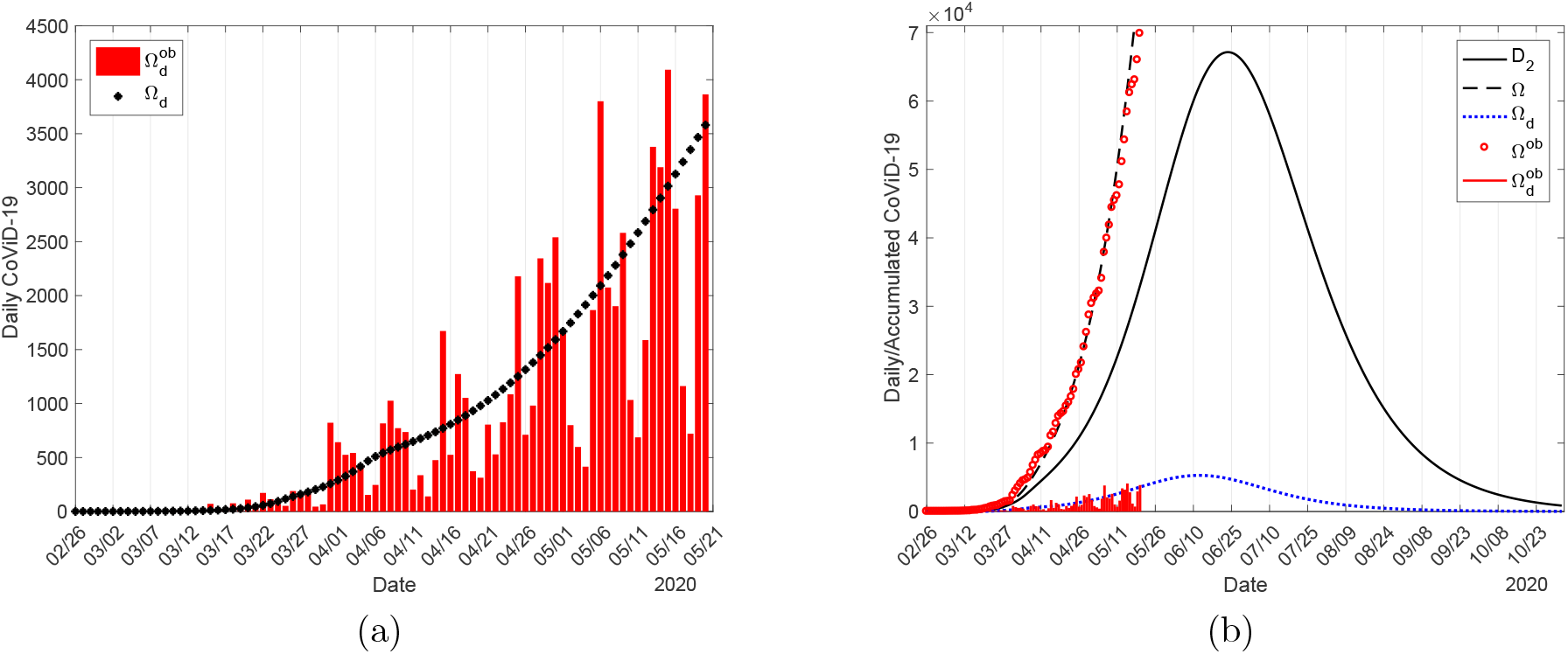
The calculated curve Ω_*d*_ and observed daily cases in São Paulo State (a), and the initial part of the estimated curve Ω with observed data Ω^*ob*^, the extended Ω_*d*_ and daily observed cases 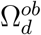, and severe covid-19 cases *D*_2_ (b).

On June 12, the peak of the daily cases of covid-19 predicted by the model reaches 5, 286. On June 23, the peak of *D*_2_ estimated by the model is 67, 140, and the number of accumulated cases Ω is 243, 000, which is 362% of the peak of *D*_2_, and 63% of cases when the first wave of epidemic ends (386, 700). These values provided by the model correspond to the epidemic with isolation without release.

### Covid-19 in Spain – Lockdown

Spain has 47.4 million inhabitants [19] with 25.8% of elder population [20], and the demographic density is 92.3*/km*^2^ [12]. In Spain, the first confirmed case of covid-19 occurred on January 31, 2020. However, the daily registering of covid-19 began on February 20 (3 cases), the first 28 deaths were registered on March 8 when reached 1, 535 cases, and on March 16 lockdown was implemented. We evaluate the model parameters using daily collected data (see Figure 19) and describe the epidemiological scenario with the lockdown.

#### Parameters evaluation

Using data from January 31 to May 20 collected from Spain, we evaluate the transmission (*β*_*y*_ and *β*_*o*_) rates, the proportion in the isolated populations (*k*), reduction in the transmission rates by protective measures adopted by population (*ε*) and social distancing (*ω*), and the additional mortality rates (*α*_*y*_ and *α*_*o*_)

#### Natural epidemic – Evaluating the transmission rates

We evaluate the transmission rates during the natural epidemic taking into account the confirmed cases from January 31 (*t*_1_) to March 21 (*t*_51_), and using equation (33). The evaluated values are *β*_*y*_ = 0.67 and *β*_*o*_ = 0.74 (both in *days*^*−*1^), where *ψ* = 1.1, resulting in the basic reproduction number *R*_0_ = 8.0 (partials *R*_0*y*_ = 5.81 and *R*_0*o*_ = 2.19), according to equation (30). Figure 9 shows the estimated curve of Ω and the observed data (a), and the extended curves for young Ω_*y*_, elder Ω_*o*_, and total Ω persons (b). On June 30, the accumulated cases for young Ω_*y*_ elder Ω_*o*_ persons approach plateaus, assuming, respectively, 564, 000, 611, 000, and 1, 175, 000.

**Figure 9:**
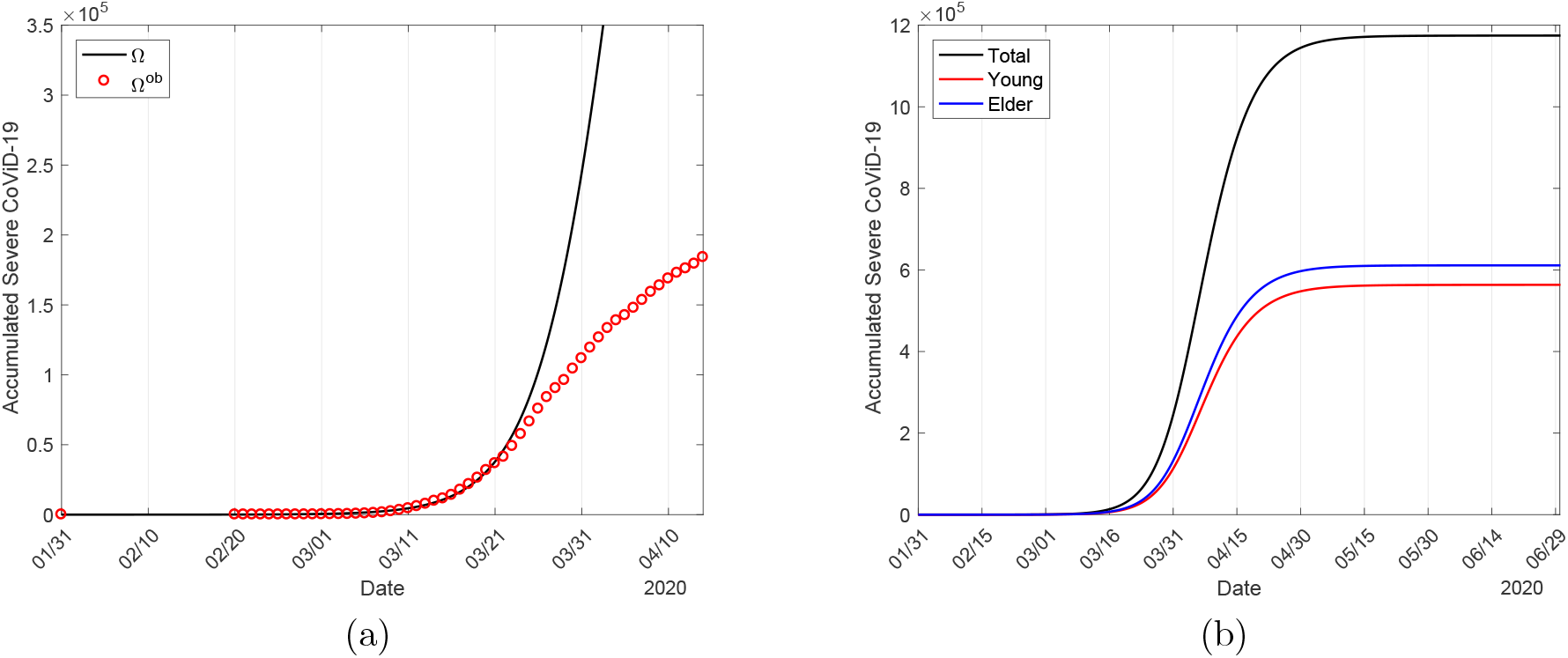
The estimated curve of the severe covid-19 cases Ω for the natural epidemic and observed data in Spain (a), and the extended curves (b).

The estimated basic reproduction number *R*_0_ in Spain is lower than São Paulo State. The inhabitants in Spain is 6% higher, but the demographic density is 48% lower than those in São Paulo State. This decrease by almost half in the demographic density may explain the 13% lower in the basic reproduction number. The elder population in Spain is 40% higher than São Paulo State, and the evaluated *ψ* is 4.4% lower than that estimated for São Paulo State.

#### Transition epidemic – Evaluating the reduction in the transmission rates in circulating and locked-down populations

On March 16, the lockdown was implemented in Spain. But the data described in Materials and Methods showed a period (from March 22 to 28) of transition from natural epidemic to lockdown effectively reducing the epidemic. Hence, we must consider covid-19 transmission in circulating and locked-down populations.

Initially, following the procedure adopted in São Paulo State, we vary the proportion of the population in lockdown, *k* = 0.5, 0.6, 0.7, 0.8, and 0.9, and evaluate the epidemic in the circulating population. Notice that the epidemic occurring in the circulating population must be lower than the observed cases of severe covid-19 if the infection is also occurring in the locked-down population. Figure 10(a) below shows that, for the proportions 0.8 and 0.9 in lockdown, the occurrence of infections in the circulating population is lower than the observed data in Spain, but *k* = 0.7 shows the number of cases provided by the model following sigmoid-shape and increasing beyond the observed data.

**Figure 10:**
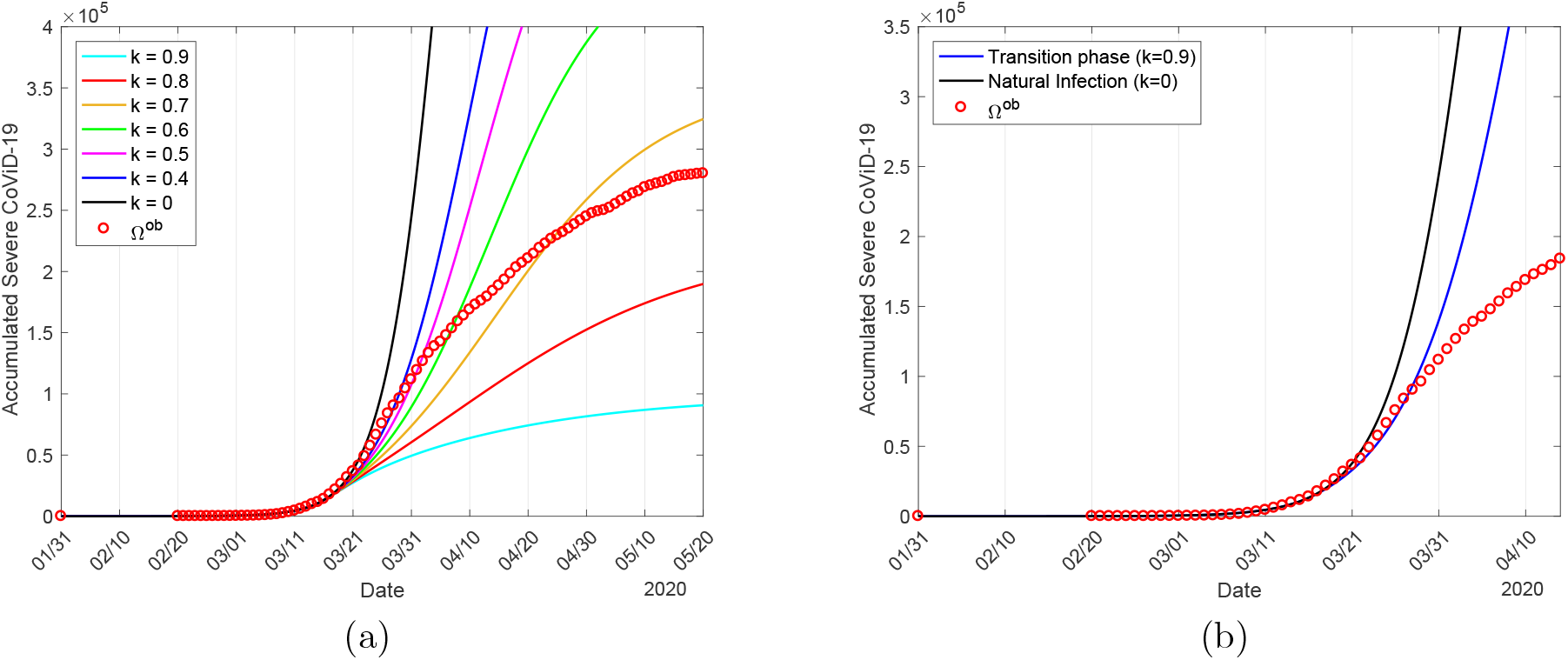
The curves of Ω for the proportions in isolation in Spain *k* = 0, 0.4, 0.5, 0.6, 0.7, 0.8 and 0.9, and observed data (a), and the estimated curves of Ω for natural and transition epidemic considering *k* = 0.9 in population in lockdown (b).

Hence, we chose *k* = 0.9 to represent the lockdown in Spain, instead of 80%, and evaluate the epidemic in circulating and locked-down populations by fixing the transmission rates *β*_*y*_ = 0.67 and *β*_*o*_ = 0.74 (both in *days*^*−*1^). We consider that during the transition of the epidemic, the populations in circulation (*k* = 0.1) and lockdown (*k* = 0.9) did not adopt any protective measure (*ε* = 1), but the population in lockdown has restricted contact (*ω >* 1). Hence, taking into account the confirmed cases from March 22 (*t*_1_) to 28 (*t*_7_) and using equation (33), the evaluated decreasing factor in the population in lockdown is *ω* = 1.5, resulting in 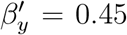 and 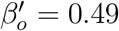 (both in *days*^*−*1^). Figure 10 shows the curves of Ω for *k* = 0.5, 0.6, 0.7, 0.8, and 0.9 and the observed data (a), and the curves of Ω for the natural epidemic and epidemic in the transition phase (b).

When a lockdown is implemented during the epidemic, those who are harboring the virus can be found in the isolated population. At the time of the beginning of lockdown on March 16, we have

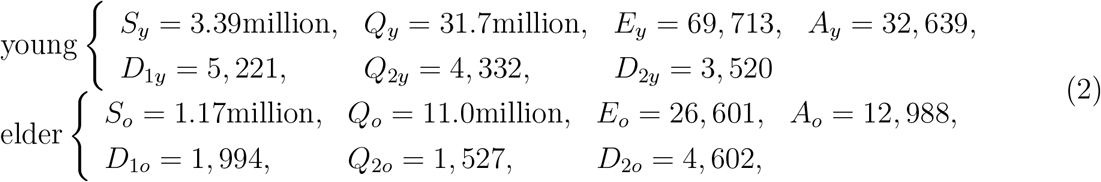

with *Q*_1*y*_ = *Q*_1*o*_ = *Q*_3*y*_ = *Q*_3*o*_ = 0 and *I* = 188, 430. When 90% (*k* = 0.9) of the infectious persons in each class is transferred to the isolated classes, the total number of isolated persons harboring virus 134, 240 can trigger a new epidemic in the population in lockdown with an elevated number of susceptible persons. When a new epidemic begins in the population in lockdown, the classes *Q*_*y*_ and *Q*_*o*_ become *S*_*y*_ and *S*_*o*_.

#### Epidemic with lockdown – Evaluating the proportion of the population in lockdown and reduction in the transmission rates

Lockdown was implemented on March 16, but the actual effects began after the transition period. We fix the previously estimated transmission rates *β*_*y*_ = 0.67 and *β*_*o*_ = 0.74 (both in *days*^*−*1^) in the natural epidemic, and the reduction in the transmission rate *ω* = 1.5 in the population in lockdown (*k* = 0.9) in the epidemic in the transition period. The parameters *ε* and *ω* corresponding to the third period of the epidemic are evaluated taking into account the confirmed cases from March 24 (*t*_1_) to May 20 (*t*_57_), and using equation (33).

We evaluate the changes in the protective measures (*ε*) in the circulating population and the decrease in the transmission rates (*ω*) in the locked-down population. In the circulating population, the evaluated protective factor is *ε* = 0.5, resulting in 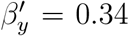 and 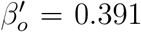 (both in *days*^*−*1^), while in the population in lockdown, the evaluated decreasing factor is *ω* = 11.5, resulting in 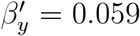 and 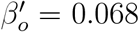 (both in *days*^*−*1^). Figure 11 shows the estimated curves of Ω and the observed data (a), and the illustration of the curves of *D*_2_ occurring in circulating (continuous) and locked-down (dashed) populations (b). The estimated curves Ω_*y*_, Ω_*o*_, and Ω = Ω_*y*_ +Ω_*o*_ approach plateaus, and on October 7, the values are, respectively, 147, 100, 173, 100, and 320, 200.

**Figure 11:**
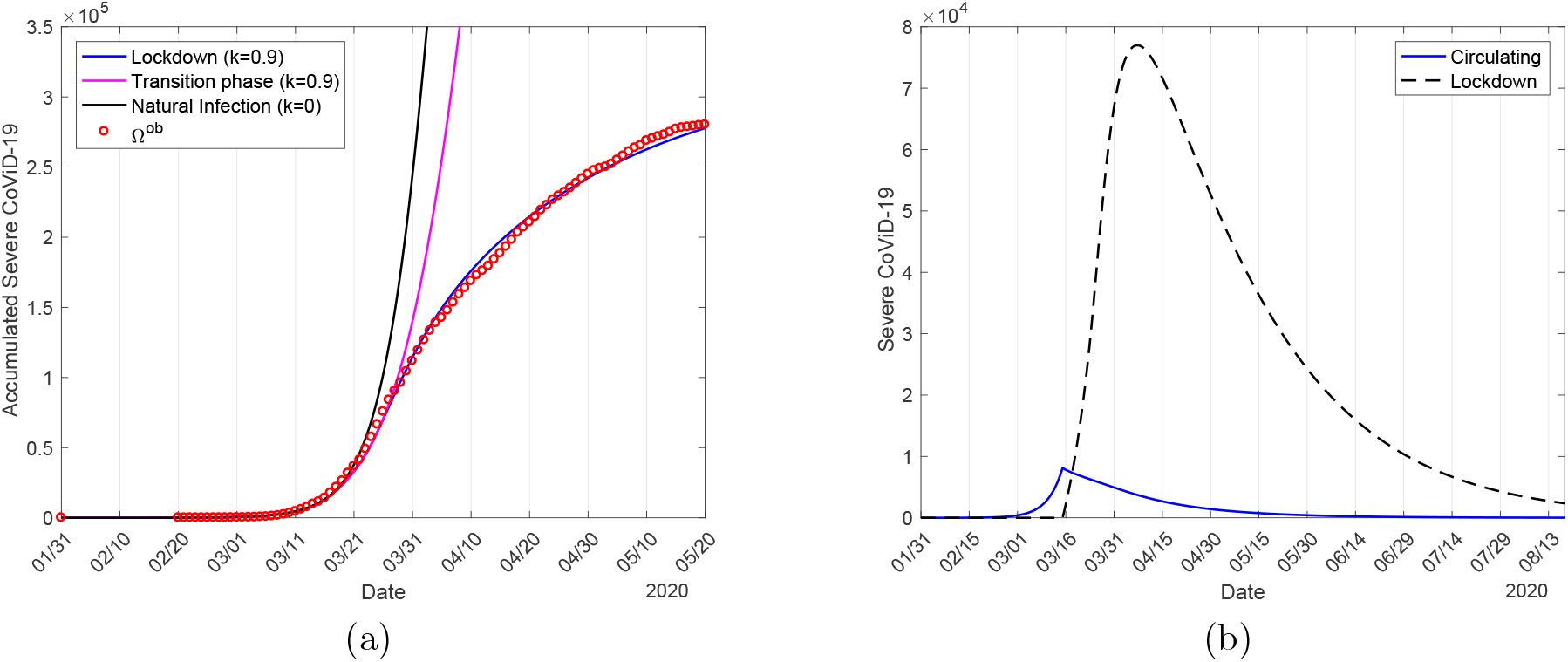
The estimated curves of Ω for natural epidemic, epidemic in transition phase, and epidemic in lockdown and observed data in Spain (a), and severe covid-19 cases in circulating and locked-down population (b). The flattened curve in (a) is the sum of two curves in (b).

For *k* = 0.8, better evaluations in *ε* and *ω* were obtained considering the effectiveness of lockdown appearing on March 25, which is exactly 9 days after March 16. The estimated protective factor in the circulating population is *ε* = 0.45, resulting in 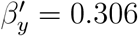 and 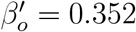 (both in *days*^*−*1^), and for the locked-down population, the decreasing factor in the transmission rates is *ω* = 12.3, resulting in 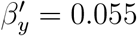 and 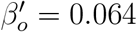 (both in *days*^*−*1^).

#### Evaluating the additional mortality rates

Taking into account confirmed deaths from March 8 (*t*_1_) to May 20 (*t*_73_) and using equation (34), we evaluate the additional mortality rates *α*_*y*_ = Γ*α*_0_ and *α*_*o*_. To evaluate the mortality rates, we fix the previously estimated transmission rates *β*_*y*_ = 0.67 and *β*_*o*_ = 0.74 (both in *days*^*−*1^) in the natural epidemic, the reduction in the transmission rate *ω* = 1.5 in the population in lockdown (*k* = 0.9) in the epidemic during the transition, and when lockdown effectively affects epidemic, protection factor *ε* = 0.5 in the population in circulation and reduction in the transmission *ω* = 11.5 in the locked-down population.

We consider Δ = 15 *days* and let Γ = 0.05 in Spain (95.5% of deaths are occurring in the elder persons with severe covid-19 [21]), and the evaluated additional mortality rates are *α*_*y*_ = 0.00273 and *α*_*o*_ = 0.0105 (both in *days*^*−*1^). Figure 12 shows the estimated curve of Π, from equation (16), and the observed death data (a), and the extended curves of the number of covid-19 deaths for young Π_*y*_, elder Π_*o*_, and total Π = Π_*y*_ + Π_*o*_ persons (b). The estimated curves Π_*y*_, Π_*o*_, and Π approach plateaus, and on October 7, the values are, respectively, 920 (0.6%), 31, 230 (18%), and 32, 150 (10%). The percentage between parentheses is the severe covid-19 case fatality rate Π*/*Ω.

**Figure 12:**
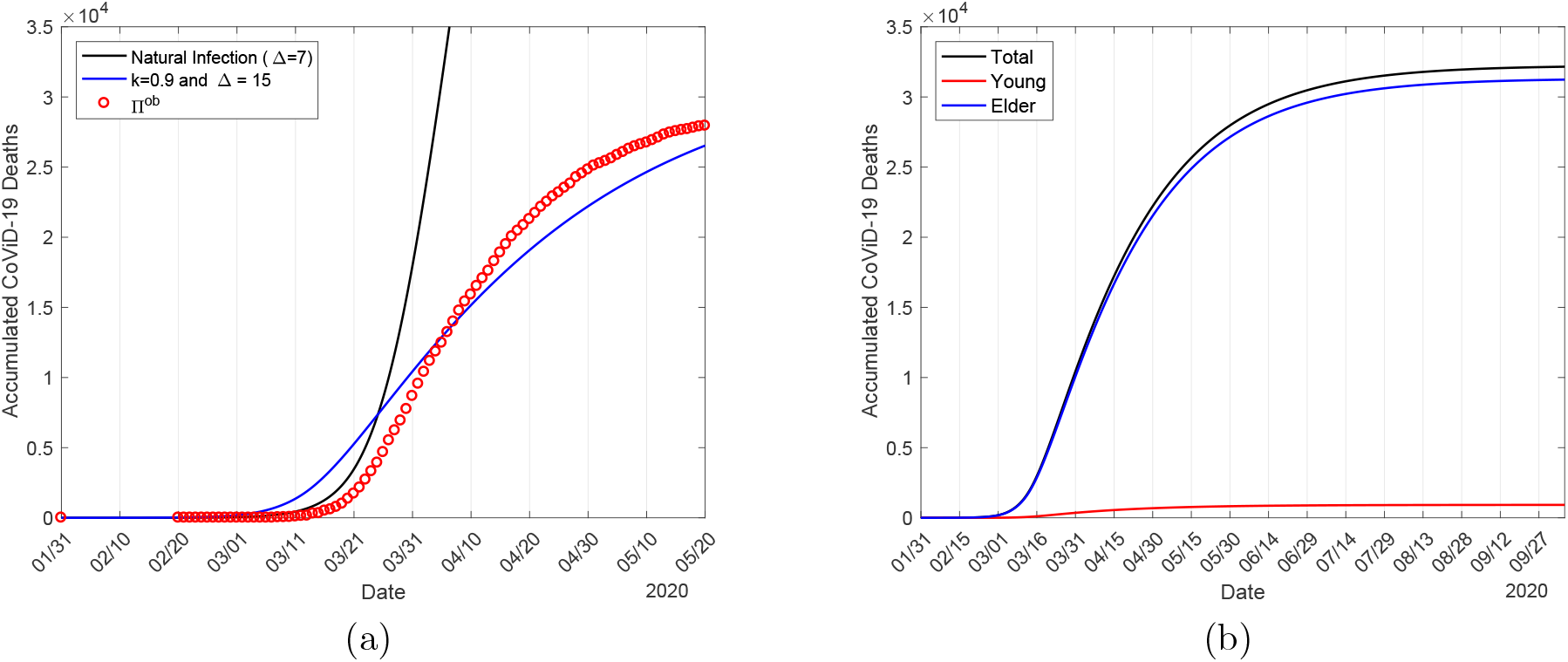
The estimated curve of accumulated deaths due to covid-19 Π and the observed data in Spain (a), and the extended curves for young Π_*y*_, elder Π_*o*_, and total Π = Π_*y*_ + Π_*o*_ persons (b).

At the end of the first wave of the epidemic, 97% of all deaths occurred in elder subpopulation. The number of the new cases of covid-19, from equation (13), for young Φ_*y*_, elder Φ_*o*_, and total Φ = Φ_*y*_ + Φ_*o*_ persons are, respectively, 9.21 million, 3.47 million, and 12.68 million. The infection fatality rate (Π*/*Φ) in young, elder and all persons are, respectively, 0.01%, 0.9%, and 0.25%.

#### Epidemiological scenario with lockdown

In Figure 13, we show the effects of interventions on the dynamic of the new coronavirus. During the three phases of the epidemic (natural, in transition, and effective lockdown), we observe decreasing in the peaks of severe covid-19 *D*_2_, which move to the right. Figure 13(a) shows the curves representing the natural epidemic, epidemic in transition, and epidemic in effective lockdown. In Figure 13(b), we show the number of immune persons *I* corresponding to the three cases shown in Figure 13(a). The curves follow a sigmoid-shape.

**Figure 13:**
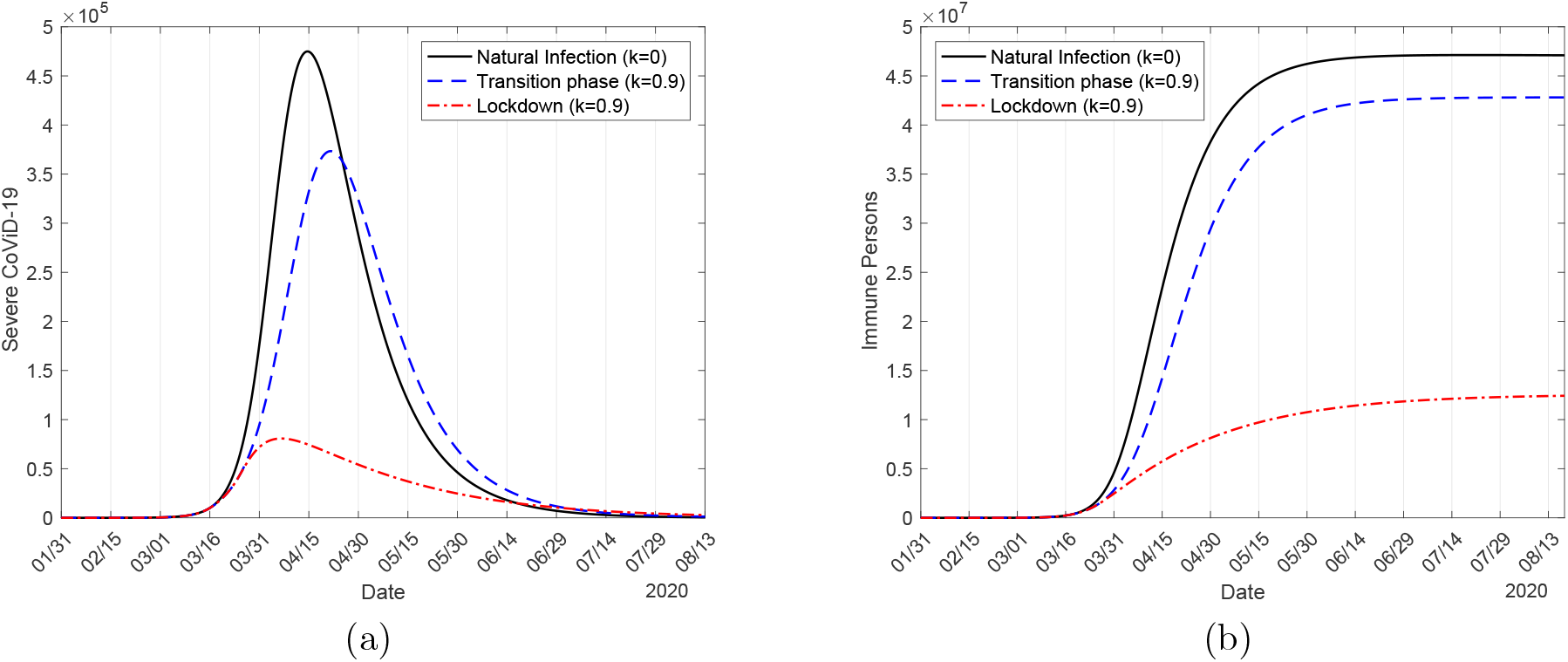
The curves of the natural epidemic (*k* = 0), the epidemic in the transition phase (*k* = 0.9), and the epidemic with lockdown (*k* = 0.9) (a), and the number of immune persons *I* (b).

In the natural epidemic, the numbers of immune persons *I*_*y*_, *I*_*o*_, and *I* increase from zero to, respectively, 35 million, 12 million, and 47 million on June 15. In the epidemic with lockdown, the numbers are, on June 15, 8.37 million (24%), 3.09 million (25.75%), and 11.46 million (24.38%). Figure 13(b) shows only *I* with and without interventions. The percentage between parentheses is the ratio between with and without interventions *I*(*k, ε*)*/I*(0, 1) on June 15.

Let us compare the peak of *D*_2_. The peaks for young, elder and total persons in the natural epidemic are, respectively, 202, 600, 272, 700, and 475, 300, occurring on April 13, 15, and 14. In the epidemic with lockdown, the peaks for young, elder and total persons are, respectively, 32, 850 (16%), 48, 190 (18%), and 80, 750 (17%), which occurred on April 5, 7 and 6. The percentage between parentheses is the ratio between natural epidemic and epidemic with lockdown *D*_2_(*k, ε*)*/D*_2_(0, 1).

Figure 14 shows the effective reproduction number *R*_*ef*_ and *D*_2_ during the epidemic in circulating (a) and locked-down (b) populations. To be fitted together in the same frame with *R*_*ef*_, the curve of *D*_2_ is divided by 1, 000 (a) and 12, 000 (b).

**Figure 14:**
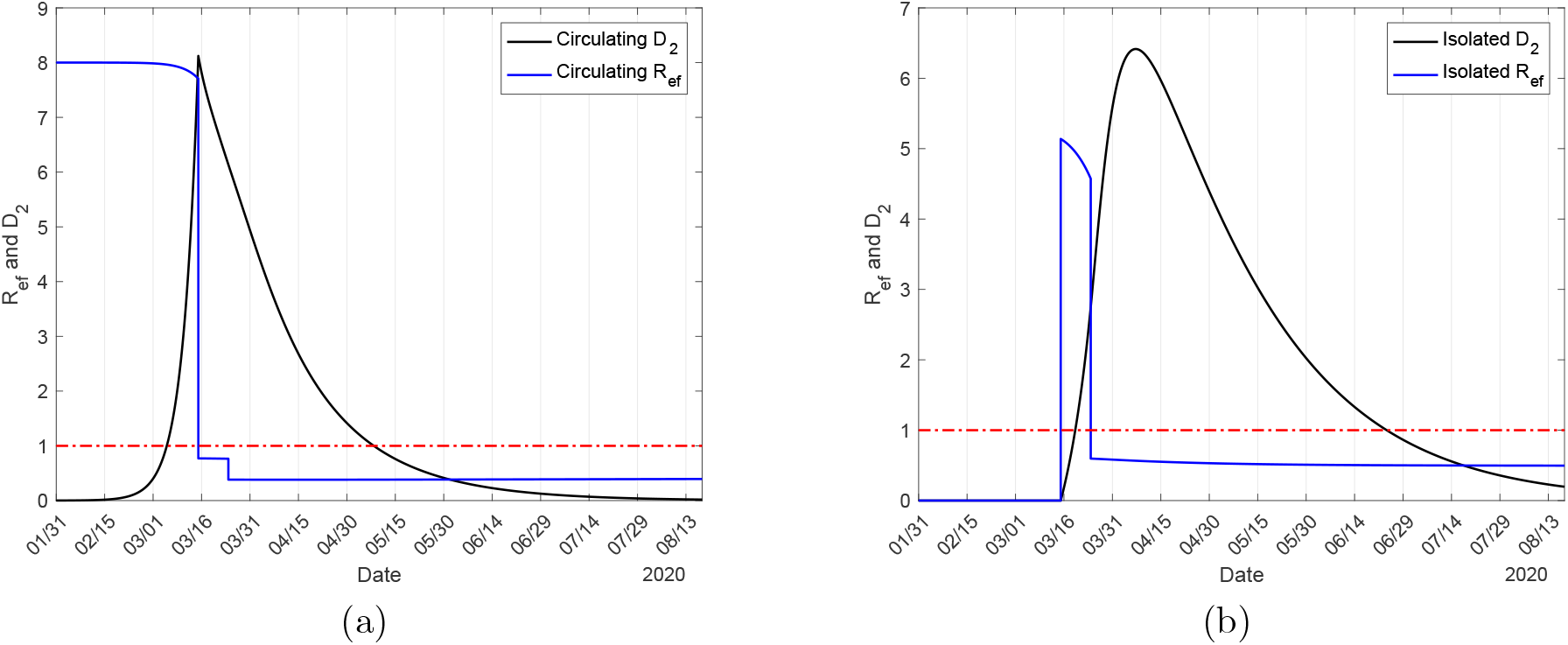
The effective reproduction number *R*_*ef*_ for epidemic with lockdown in circulating (a) and locked-down (b) population in Spain. The number of severe covid-19 cases *D*_2_ must be multiplied by 1, 000 (a) and 12, 000 (b).

For 90% of the population in lockdown in Spain, the basic reproduction number *R*_0_ = 8.0 decreased to *R*_*ef*_ = 0.771 and *R*_*ef*_ = 5.14, respectively, in circulating and locked-down populations on March 16. During a short period of transition from natural to lockdown epidemic, the high effective reproduction number in the 90% in the lockdown resulted in a high number of infections (see Figure 11(b), practically all cases are originated in locked-down population), and the daily covid-19 cases increased, reaching the peak on March 27. On March 24, when the effectiveness of lockdown is observed, we observe another reduction in the effective reproduction number to *R*_*ef*_ = 0.382 and *R*_*ef*_ = 0.59, respectively, in circulating and locked-down populations. Although *R*_*ef*_ *<* 1, the number of new cases of covid-19 does not decrease quickly due to the high number of susceptible individuals. Hence, the example of Spain demonstrated that it is not enough to decrease the effective reproduction number below unity if the number of susceptible individuals is higher. On May 4 (phase 0 of release in Spain [22]) and June 8 (phase 3 of release) the effective reproduction number assumes, respectively, 0.53 and 0.51.

Figure 15 shows the curve Ω_*d*_ derived from Ω and the observed data in Spain (a), and the estimated curve of Ω with observed data, the curves *D*_2_ and Ω_*d*_ with the observed data (b).

**Figure 15:**
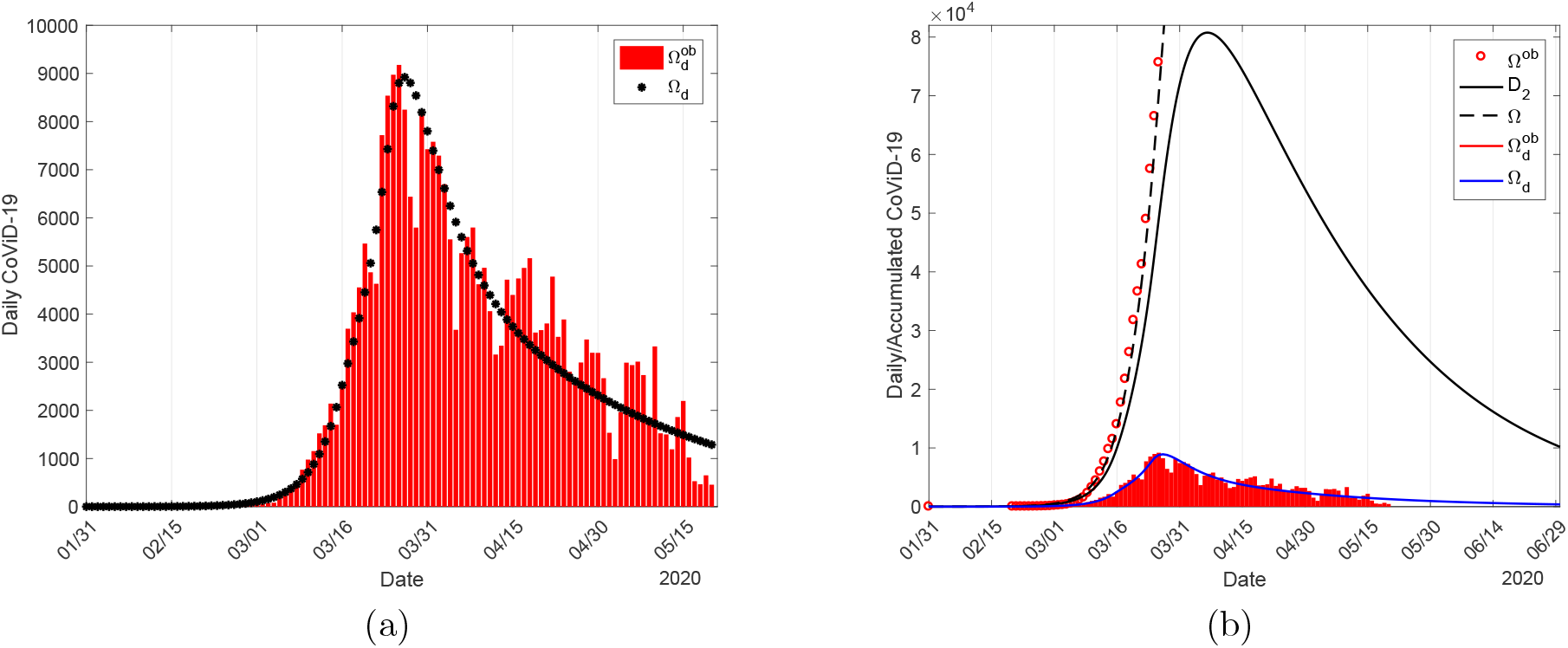
The calculated curve Ω_*d*_ and the observed daily cases in Spain (a), and the initial part of the estimated curve Ω with observed data Ω^*ob*^, the extended Ω_*d*_ and daily observed cases 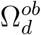, and severe covid-19 cases *D*_2_ (b). All curves are the sum of the cases in circulating and locked-down populations.

The peaks of the estimated curves Ω_*d*_ and *D*_2_ are, respectively, 8, 922 and 80, 750, which occur on March 27 and April 6. The peak of estimated daily cases is 97.2% (2.8% difference) and occurred one day later in comparison with the peak of observed daily cases, which is 9, 177 occurring on March 26. However, for *k* = 0.8, the peaks of the estimated curves Ω_*d*_ and *D*_2_ are, respectively, 9, 566 and 80, 020 occurring on March 28 and April 6. The peak of estimated daily cases is 104.2% (4.2% difference) and occurred two days later in comparison with the peak of observed daily cases. Hence, 90% of the population in lockdown explains better the daily observed data in Spain than 80%.

### Comparing covid-19 epidemics in São Paulo State and Spain

The widespread epidemic of covid-19 led Spain to implement lockdown, and the number of asymptomatic persons was higher as shown in equation (2), while São Paulo State isolated population earlier, and the number of asymptomatic persons was not so high as shown in equation (1). In the population in lockdown in Spain, the numbers of infectious and susceptible persons are, respectively, 3 and 1.8-time more than those found in the isolation in São Paulo State. The number of new cases of infection is proportional to the product of the numbers of infectious and susceptible persons, hence the population in lockdown in Spain has 5.4-time more risk of an outbreak of the epidemic than in São Paulo State. For this reason, we neglected covid-19 transmission in the isolated population in São Paulo State, however, to explain the observed data in Spain, we allowed a low transmission (through restricted contact occurring in the household and/or neighborhood) in the locked-down population. Indeed, the model provided that almost all cases of severe covid-19 cases were originated from the locked-down population in Spain, although *R*_*ef*_ = 0.59. However, the epidemic of covid-19 in São Paulo State occurred in the circulating population, with the effective reproduction number jumping down to *R*_*ef*_ = 4.35 when the isolation in 53% of the population was implemented on March 24, which decreased more to *R*_*ef*_ = 2.1 with the adoption of protective measures on April 4. According to the definition in [7], São Paulo State is an example of mitigation, while Spain, suppression.

The elapsed time from the first case to the adoption of lockdown in Spain was 45 days, while isolation in São Paulo State, 27 days, a difference of 18 days. The value of *R*_*ef*_, from the beginning of the epidemic to the implementation of lockdown/isolation, decreased by 0.29 (with 8, 122 cases of severe covid-19) and 0.02 (with 397 cases), respectively, in Spain and São Paulo State. Moreover, the peak of severe covid-19 cases occurred on April 6 (66 days after the beginning of the epidemic) with 80, 750 cases in Spain, which is 17% of 474, 900 cases in the natural epidemic. In São Paulo State, the estimated peak of severe cases will occur on June 23 (118 days after the beginning of the epidemic) with 67, 140 cases (83% of Spain), which is 18% of 386, 400 cases in the natural epidemic. The 52 days gained by São Paulo State were extremely valuable to avoid the overloading in the health system, sowing that the early adoption of isolation or lockdown is essential to control the epidemic.

The estimated *R*_0_ for covid-19 in Spain is 87% of that in São Paulo State, although the peak and the accumulated cases at the end of the first wave of the natural epidemic are, respectively, 123% and 124% of those in São Paulo State. This finding can be understood by analyzing the partial reproduction numbers *R*_0*y*_ for young and *R*_0*o*_ for elder subpopulations. We estimated, for São Paulo State, *R*_0*y*_ = 7.73 and *R*_0*o*_ = 1.51. From the model, the fractions of young and elder susceptible persons reach, in the long-term epidemic, respectively, 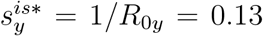 and 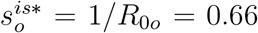. However, when young and elder subpopulations are not separated, but are interacting in the circulating population, we obtained 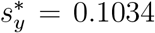 and 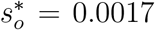. Notice that the difference 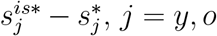, is the additional proportion of susceptible persons infected due to interaction, being 2.7% for young and 66% for elder persons, showing that elder persons are 24-time more risk than young persons when they interact. For Spain, we estimated *R*_0*y*_ = 5.81 and *R*_0*o*_ = 2.19, which are 75% and 145% of those estimated in São Paulo State. For the number of accumulated cases, Spain has 93% (young) and 179% (elder) of severe covid-19 cases of those in São Paulo State. For the number of cases at the peak of the epidemic, Spain has 90% (young) and 168% (elder) of those in São Paulo State. Finally, in Spain, the majority of infections were occurring in 90% of the locked-down population with 25.8% of the elder population, while in São Paulo State, the infections were occurring in 47% of the isolated population with 15.3% of the elder population. Therefore, the higher number of cases with lower *R*_0_ in Spain can be explained epidemiologically by lockdown, which allowed a higher number of elder persons in close contact with young persons, increasing the infection in the vulnerable elder subpopulation.

Both estimated curve of Ω and the observed data in Spain after the implementation of lockdown follow the shape of *R*_*ef*_ lower than 1 (Figures 11(a) and 19(b)), while the curve of Ω and observed data after the adoption of isolation in São Paulo follow the shape of *R*_*ef*_ higher than 1 (Figures 3(a) and 17(b)). Briefly, during the epidemic, the shape of the curve of accumulated cases of severe covid-19 Ω is sigmoid, that is, presents a quick increase in the first phase (*R*_*ef*_ *>* 1, upward concavity) followed by a slow increase (*R*_*ef*_ *<* 1, downward concavity). At the end of the first wave of the epidemic, the accumulated severe covid-19 cases in Spain is 320, 200, and in São Paulo State, 386, 600 (121% of the cases in Spain), although the peak of the epidemic in Spain is higher. As a consequence, the lockdown implemented in Spain reduced the number of covid-19 cases (27% of the natural epidemic) more than isolation adopted in São Paulo State (41% of the natural epidemic), which impacts on the number of immune persons. The number of young and elder immune persons are, respectively, 15.14 million (99%) and 2.82 million (97%) for São Paulo State, and 9.13 million (99%) and 3.41 million (98%) for Spain. The percentage between parentheses is the ratio of the numbers of immune persons and new cases of covid-19 *I/*Φ. In São Paulo State, the total number of immune persons at the end of the first wave of the epidemic is 40% of the population, while in Spain, 26.5%. On June 15, the proportion of the immune person in São Paulo State is 16.7%. In Spain, on May 4, June 8 and 15, the proportions of the immune persons are, respectively, 18%, 23.5%, and 24.4%, showing that on June 8 Spain is close to the end of the first wave of the epidemic. However, if lockdown/isolation and protective measures are relaxed or abandoned, Spain and São Paulo State will be under higher risk to trigger a second wave of the epidemic due to a high number of susceptible persons and a low number of immune persons at the end of the first wave of the epidemic.

At the end of the first wave of the epidemic, the estimated number of deaths in Spain is 32, 150 (10% of total cases), while in São Paulo State is 23, 780 (6% of total cases), which is 74% of the total deaths in Spain. The severity case fatality rate in São Paulo State is 3.7-time higher in young, and 1.4-time lower in elder in comparison with Spain. Both severity case and infection fatality rates for Spain for young and elder persons are, respectively, around 30% and 138% of the rates obtained for São Paulo State. These rates in elder subpopulation could be explained by the life expectancy (in São Paulo State is 78.4 *years* and in Spain, 83.4 *years*), because 59.7% of deaths occurred in elder persons with 80 years old or more in Spain [20]. However, a higher proportion of uncontrolled comorbidity in the young subpopulation in São Paulo State could increase the number of deaths [23].

On May 20, the number of deaths was around 12% of severe covid-19 cases in Spain, while in São Paulo State, it was around 7%. The number of deaths is closely related to the number of severe covid-19 cases. As we pointed out, the close interaction between locked-down young and elder subpopulations (the presence of infectious young individuals increases the risk of infection among elders [15]) increased the epidemic in the elder subpopulation, increasing deaths. Moreover, the quick increase in the number of severe covid-19 in Spain overloaded hospitals and, hence, may have contributed to an increase in the death of untreated patients, especially elders. The current relatively low number of deaths in São Paulo State compared with that observed in Spain indicate that the health care system must be prepared to avoid the overloading in hospitals.

The lockdown adopted in Spain was an extreme measure imposed by the critical epidemiological situation. To avoid the peak of the forthcoming epidemic, São Paulo State cogitated to adopt lockdown besides the isolation already implemented. The implementation of lockdown in an ascending phase of the epidemic will transfer an elevated number of asymptomatic persons to isolation, permitting close contact with the population already in isolation (see [15]). Hence, the adoption of lockdown in a population already in isolation may trigger a new epidemic, especially in the case of the SARS-CoV-2 with high transmissibility (higher *R*_0_). As we pointed out, Spain showed that the combination of higher numbers of infectious and susceptible persons in the locked-down population allowed a persistent low transmission even when *R*_*ef*_ is lower than one.

### Remarks

In the literature, the usually assumed basic reproduction number *R*_0_ is around 3, see for instance [6] and [7]. However, Li *et al*. [8] explicitly cited that, by using data from January 10 to February 8, 2020, they estimated the effective reproduction number *R*_*ef*_, arguing that the most recent common ancestor could have occurred on November 17, 2019. The time elapsed from November 17, 2019 (the first case) to January 10, 2020 (the first day in the estimation) is 54 days. On January 23, 2020, Wuhan and other cities of Hubei province imposed lockdown. As we pointed out in Figure 1(b), taking into account the entire data, or restricting the interval of data around the implementation of isolation in São Paulo State, the estimated *R*_*ef*_ must be lower than 5.09.

From Figures 7 and 14, the effective reproduction number *R*_*ef*_ for São Paulo State, 54 days after the beginning of the epidemic, is 2.1 (April 20), while for Spain, *R*_*ef*_ = 0.6 (March 25). In other words, using covid-19 data beginning from April 20 (São Paulo State) or March 25 (Spain), probably the estimated effective reproduction number will be close to those retrieved from Figures 7 and 14. On the other hand, if we estimate the basic reproduction number using SIR model with different infective persons at *t* = 0, we obtain, using data collected from São Paulo State, *R*_0_ = 3.22 (for *I*(0) = 10), or *R*_0_ = 2.66 (for *I*(0) = 25), or *R*_0_ = 2.38 (for *I*(0) = 50), with other initial conditions being *S*(0) = 44.6 million, and *R*(0) = 0.

The reliable estimation of *R*_0_ is important because this number determines the magnitude of effort to eradicate infection. For instance, the efforts of vaccination to eradicate an infection must be vaccinating a fraction equal or greater than 1 *−* 1*/R*_0_ of susceptibles [9]. In [14], analyzing vaccination as a control mechanism, if *R*_0_ is reduced by the vaccine to value lower than one, the number of cases decreased following exponential-type decay, as we observed in Figure 14 describing the lockdown in Spain. Hence, instead of a vaccine, let us consider lockdown to control covid-19 transmission. If *R*_0_ = 3, we must isolate at least 67% of the population, while for *R*_0_ = 8, at least 87% of the population. As we have pointed out, 70% of the population in lockdown does not explain the covid-19 data in Spain, but 90% of the population in lockdown explained better the observed data. Hence, our estimations of *R*_0_ for São Paulo State (9.24) and Spain (8.0) using the first period of covid-19 data represent the natural epidemic. Moreover, the estimation of *R*_0_ based on the first period is in concordance with the definition of the basic reproduction number: Completely susceptible population without constraints (interventions) [9].

In [24] and [10] we estimated the additional mortality rates based on the observed data and concluded that their values fitted well at the beginning of epidemic but did not provide reliable fitting in the long-term epidemic. For instance, that method of estimation pairing the numbers of new cases and deaths at the registering time resulted in deaths of 30% up to 80% of severe covid-19 cases at the end of the first wave of the epidemic. For this reason, we had adopted a second method of estimation considering that the accumulated deaths in the elder subpopulation at the end of the first wave of the epidemic must be around 10%, underestimating the number of deaths at the beginning of the epidemic. Here, we improved the estimation method by pairing the numbers of new cases and deaths not at the registering time, but delayed in 15 days, which is suggested by comparing daily registered data of new cases and deaths, see Figure 17. Figures 4 and 12 showed that this novel method of estimation fits relatively well during the three periods of the epidemic with Δ = 15 *days*. However, we can vary Δ according to the period of the epidemic to obtain a better fitting. For instance, in Figure 12, the accumulated data of deaths corresponding to the natural epidemic is well fitted using Δ = 7 *days*.

The concept of herd immunity is associated with the protection provided by immunized persons to a specific subpopulation under a higher risk of death caused by a syndrome or co-morbidity. For instance, in the rubella infection, mass vaccination was planned to diminish the infection among pregnant women to reduce the number of congenital rubella syndrome [25]. The misunderstanding of the concept of herd immunity, and the rapid dissemination of under-estimated *R*_0_ around 3, may have led to misconducting public health policies. For instance, the United Kingdom (at the beginning of epidemic) and Sweden adopted the idea of immunization by natural infection, believing that the “herd immunity” would protect susceptible, especially elder subpopulation. However, the concept of herd immunity must not be understood as immunization by the circulating virus with high lethality but by the vaccine, which decreases the effective reproduction number *R*_*ef*_. The jump down in Figures 7 and 14 can be applied to vaccinating *k* proportion of the susceptible persons. However, when the isolation and protective measures are implemented in a population, the effective reproduction number *R*_*ef*_ jumps down, but this reduction in *R*_*ef*_ is temporary and lasts whenever the population maintains adherence to lockdown/isolation and protective measures. These non-pharmaceutical interventions can be understood as transitory herd protection, such as the herd immunity by vaccination (when available) protects especially the elder subpopulation under higher risk of infection and death.

## Conclusion

In the absence of effective treatment and vaccine, the adoption of lockdown at the very beginning of the epidemic is recommended to control the SARS-CoV-2 with high transmissibility and lethality. The second strategy is the implementation of isolating as São Paulo State did. In this case, in the circulating population, the epidemic curve is flattened to avoid the overloading in hospitals, and the immunization by the natural epidemic is increased. Unfortunately, the number of deaths due to covid-19 increases. The third strategy, the adoption of lockdown, is recommended when the epidemic is out of control, and Spain is an example.

In the second and third strategies to control the covid-19 epidemic, the severe covid-19 data before the adoption of isolation or lockdown are used to estimate the basic reproduction number. However, when the lockdown is implemented immediately after the outbreak of the epidemic, the estimation of the basic reproduction number may not be done directly from the observed data of severe covid-19 cases.

Isolation as well as lockdown are valuable measures to control an epidemic with high lethality. Due to the critical situation of the health care system, Spain imposed a rigid lockdown, which impacted the fast ascending phase of the epidemic by reducing *R*_*ef*_ below one. This reduction resulted in a peak of 80, 750 cases occurring 20 days after the implementation of lockdown (on March 16, Ω^*ob*^ = 14, 011 from the observed data). Convinced by the epidemiological situation in Spain, São Paulo State implemented isolation, as result, the peak of 67, 140 cases will occur after 91 days since isolation (on March 24, Ω^*ob*^ = 810 from the observed data). The relatively early implementation of the isolation in São Paulo State in somehow avoided the overloading in the health care system by flattening the epidemic curve.

The proportion of the immune persons at the end of the first wave of epidemic is 40% of the population in São Paulo State, and 26% of the population in Spain. On June 8 (phase 3 of the release in Spain [22]), the effective reproduction number was 0.51 and 23.5% of the population was immune, showing that Spain was close to the end of the first wave of the epidemic. This relatively safe epidemiological scenario was favorable to implement a carefully planned release. However, on June 15, the effective reproduction number was 0.98 but in the ascending phase of the epidemic, and 16.7% of the population was immune, showing that São Paulo State was far from the end of the epidemic. If the isolated persons are released in this unfavorable epidemiological scenario, the already intense transmission of SARS-CoV-2 may be enhanced. All these findings will help to plan strategies of release, which is left to further work.

## Materials and Methods

We present a mathematical model from which we obtain the basic reproduction number *R*_0_ and the effective reproduction number *R*_*ef*_, we describe the data of severe covid-19 cases and deaths collected from São Paulo State and Spain, and we obtain values for the model parameters.

### Mathematical model – Retrieving *R*_0_ and *R*_*ef*_

We present a mathematical model to describe the covid-19 epidemic and the calculation of the basic and effective reproduction numbers *R*_0_ and *R*_*ef*_. The model aims to assess the effects of isolation and lockdown on the epidemic of covid-19.

#### Formulation of the model

In a community where SARS-CoV-2 is circulating, the risk of infection is greater in elder than young persons, as well as elder persons are under an increased probability of being symptomatic and higher covid-19 induced mortality. Hence, the community is divided into two groups, composed of young (60 years old or less, denoted by subscript *y*), and elder (60 years old or more, denoted by subscript *o*) persons. The vital dynamic of this community is described by the per-capita rates of birth (*ϕ*) and mortality (*µ*).

For each subpopulation *j* (*j* = *y, o*), all persons are divided into nine classes: susceptible *S*_*j*_, susceptible persons who are isolated *Q*_*j*_, exposed and incubating *E*_*j*_, asymptomatic *A*_*j*_, symptomatic persons in the initial phase of covid-19 (or pre-diseased) *D*_1*j*_, pre-diseased persons caught by test and then isolated *Q*_1*j*_, symptomatic persons with severe covid-19 *D*_2*j*_, mild covid-19 *Q*_2*j*_, and mild covid-19 persons isolating themselves by educational campaign *Q*_3*j*_. However, all young and elder persons in classes *A*_*j*_, *Q*_1*j*_, *Q*_2*j*_, *Q*_3*j*_, and *D*_2*j*_ enter into the same immune class *I* (this is the 10^*th*^ class, but common to both subpopulations).

The natural history of new coronavirus infection is the same for young (*j* = *y*) and elder (*j* = *o*) subpopulations. We assume that persons in the asymptomatic (*A*_*j*_), pre-diseased (*D*_1*j*_), and a fraction *z*_*j*_ of mild covid-19 (*Q*_2*j*_) classes are transmitting the virus, and other infected classes (*Q*_1*j*_, (1 *− z*_*j*_) *Q*_2*j*_ and *D*_2*j*_) are under voluntary or forced isolation. Susceptible persons are infected according to *λ*_*j*_*S*_*j*_ (known as the mass action law [9]) and enter into class *E*_*j*_, where *λ*_*j*_ is the per-capita incidence rate (or force of infection) defined by *λ*_*j*_ = *λ* (*δ*_*jy*_ + *ψδ*_*jo*_), with *λ* being

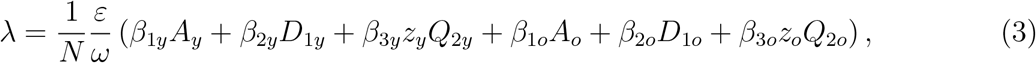

where *δ*_*ij*_ is Kronecker delta, with *δ*_*ij*_ = 1 if *i* = *j*, and 0, if *i* ≠ *j*; and *β*_1*j*_, *β*_2*j*_ and *β*_3*j*_ are the transmission rates, that is, the rates at which a virus encounters a susceptible people and infects him/her. The parameters *ε ≤* 1, and *ω ≥* 1 diminish the transmission rates. The protection factor *ε* decreases the transmission of infection by individual (face mask, hygiene, etc.) and collective (social distancing) protective measures, while the reduction factor *ω* decreases the transmission by the contact being restricted in the population in isolation (household and/or neighborhood contacts). After an average period 1*/σ*_*j*_ in class *E*_*j*_, where *σ*_*j*_ is the incubation rate, exposed persons enter into the asymptomatic class *A*_*j*_ (with probability *p*_*j*_) or pre-diseased class *D*_1*j*_ (with probability 1 *− p*_*j*_). After an average period 1*/γ*_*j*_ in class *A*_*j*_, where *γ*_*j*_ is the recovery rate of asymptomatic persons, symptomatic persons acquire immunity (recovered) and enter into immune class *I*. Possibly asymptomatic persons can manifest symptoms at the end of this period, and a fraction 1 *− χ*_*j*_ enters into mild covid-19 class *Q*_2*j*_. Another route of exit from class *A*_*j*_ is being caught by a test at a rate *η*_*j*_ and enter into class *I* (we assume that this person indeed adopts isolation, which is the reason to enter into class *I* at a rate of testing). For symptomatic persons, after an average period 1*/γ*_1*j*_ in class *D*_1*j*_, where *γ*_1*j*_ is the infection rate of pre-diseased persons, pre-diseased persons enter into severe covid-19 class *D*_2*j*_ (with probability 1 *− m*_*j*_) or class *Q*_2*j*_ (with probability *m*_*j*_), or they are caught by test at a rate *η*_1*j*_ and enter into class *Q*_1*j*_. Persons in class *D*_2_ acquire immunity after period 1*/γ*_2*j*_, where *γ*_2*j*_ is the recovery rate of severe covid-19, and enter into class *I* or die under the disease-induced (additional) mortality rate *α*_*j*_. Another route of exiting class *D*_2*j*_ is by treatment, described by the treatment rate *θ*_*j*_. Class *Q*_1*j*_ is composed of mild and severe covid-19 persons who came from class *D*_1*j*_ caught by test, hence they enter into class *D*_2*j*_ (with the rate (1 *− m*_*j*_) *γ*_1*j*_) or class *I* (with the rate *m*_*j*_*γ*_1*j*_ +*γ*_2*j*_, assuming adherence to isolation). Persons in class *Q*_2*j*_ acquire immunity after period 1*/γ*_3*j*_, where *γ*_3*j*_ is the recovery rate of mild covid-19, and enter into immune class *I*. Another route of exit from class *Q*_2*j*_ are being caught by a test at a rate *η*_2*j*_ and enter into class *I* (assumption of adherence to isolation), or enter to class *Q*_3*y*_ convinced by an education campaign at a rate *rv*_*j*_, which is temporary, hence ϖ_*j*_ is the rate of abandonment of protective measures [26].

In the model, we consider pulse isolation and intermittent (series of pulses) release of persons. We assume that there is a unique pulse in isolation at time 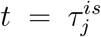, described by 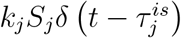, but there are *m* intermittent releases described by 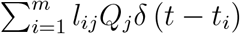, where 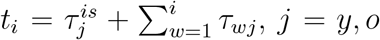, and *δ* (*x*) is Dirac delta function, that is, *δ* (*x*) = *∞*, if *x* = 0, otherwise, *δ* (*x*) = 0, with 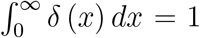. The fraction of persons in isolation is *k*_*j*_, and *l*_*ij*_, *i* = 1, 2, *…, m*, is the fraction of *i*-th release of isolated persons, with *τ*_*wj*_ being the period between successive releases.

Figure 16 shows the flowchart of the new coronavirus transmission model.

**Figure 16:**
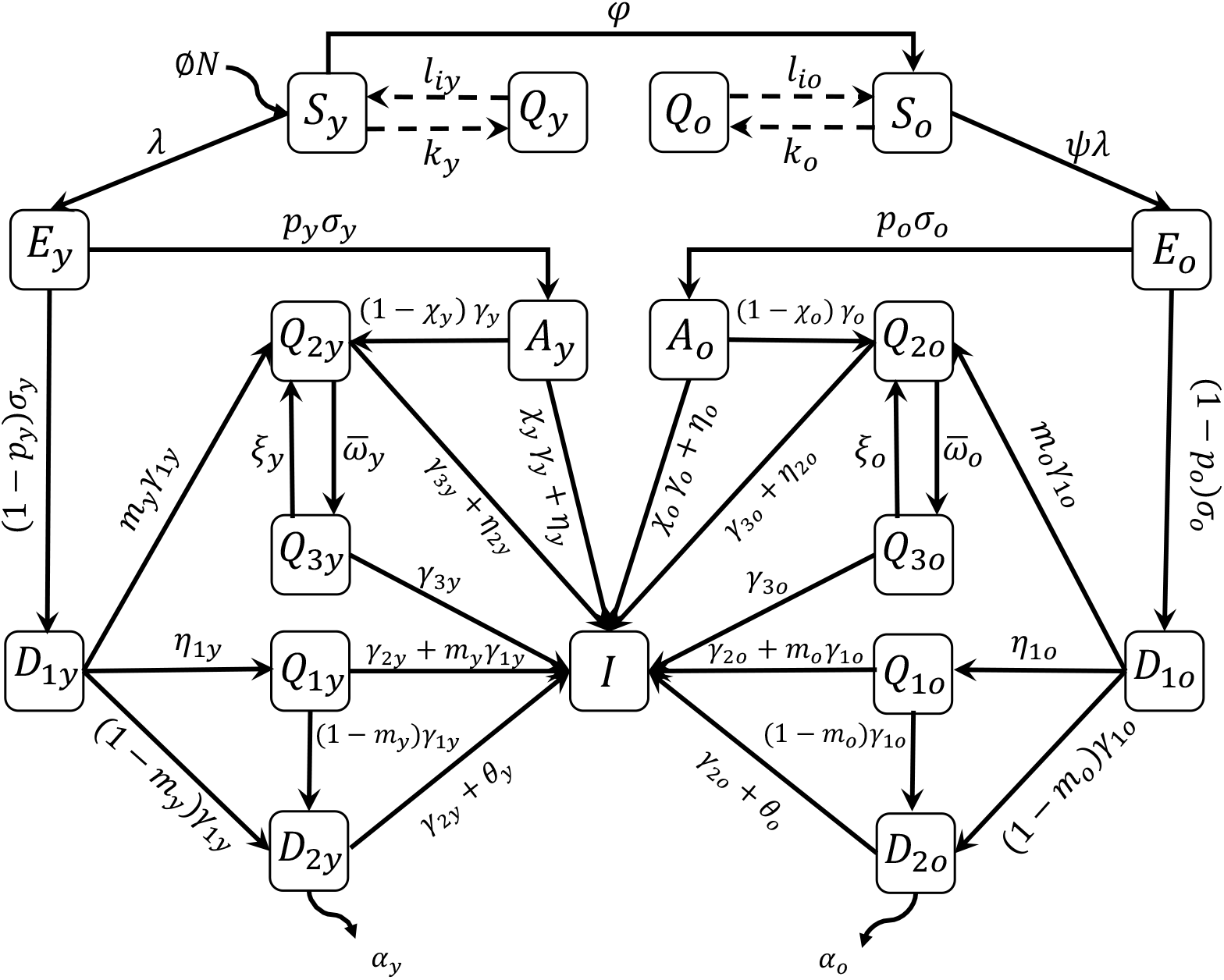
The flowchart of new coronavirus transmission model with variables and parameters.

**Figure 17:**
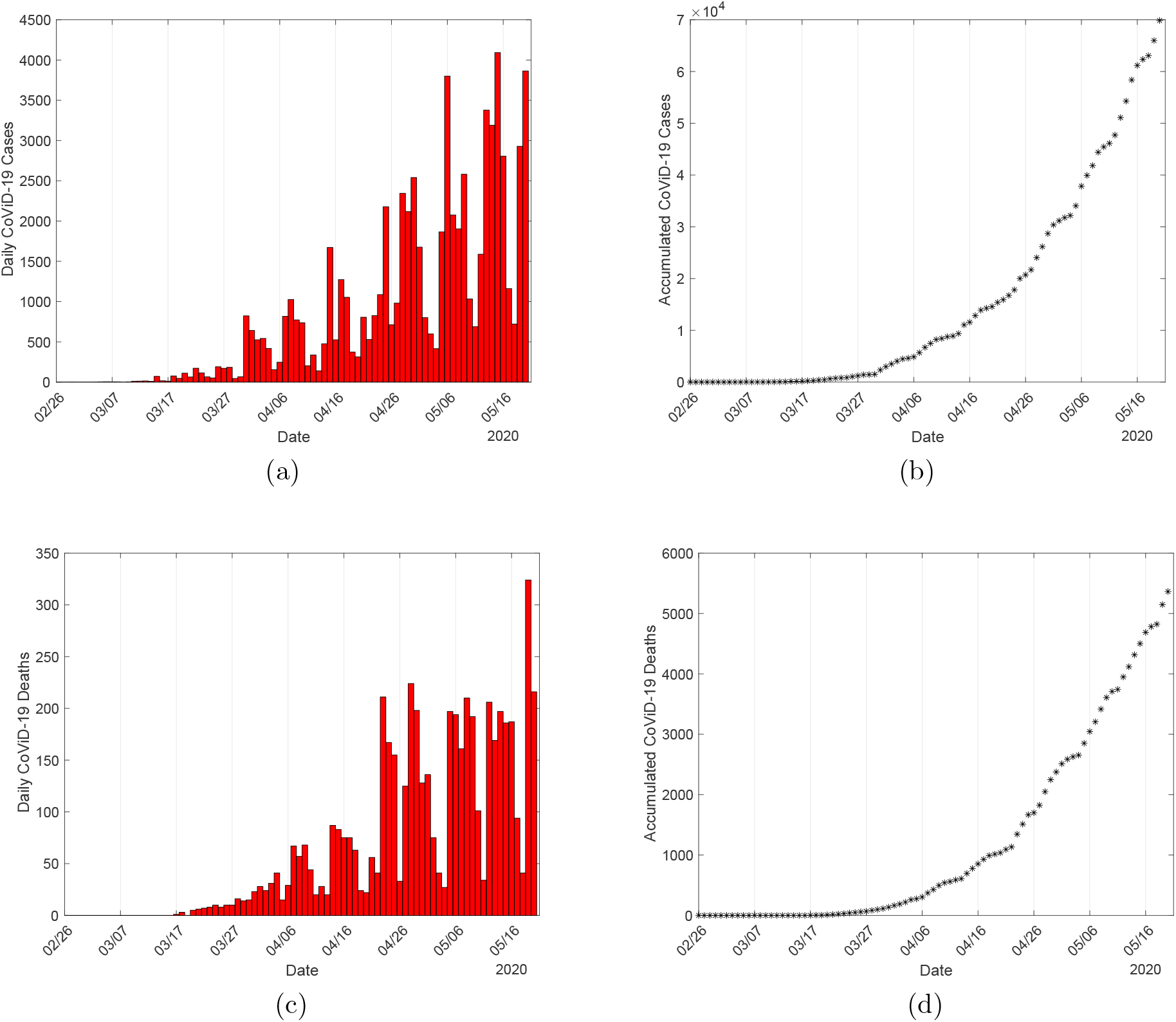
The daily (a) and accumulated (b) covid-19 cases, and daily (c) and accumulated (d) deaths, using covid-19 cases collected in São Paulo State.

**Figure 18:**
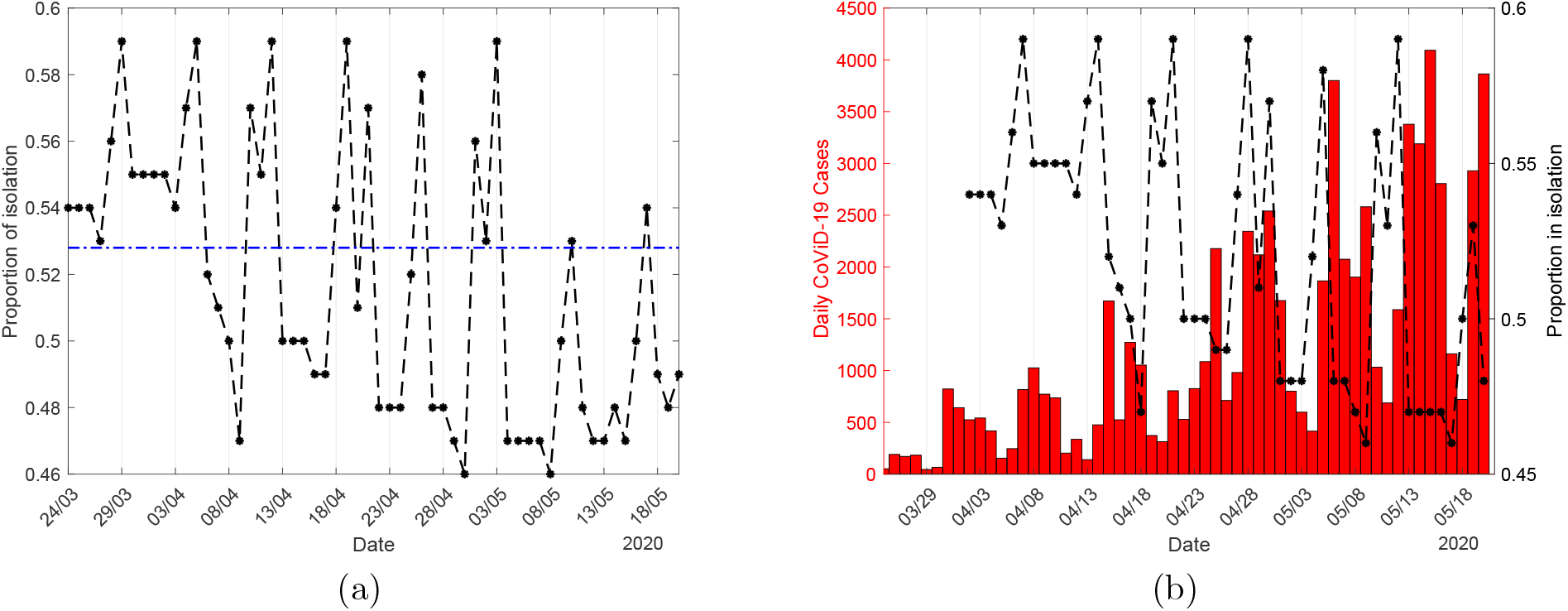
The proportion in isolation in São Paulo State (a), and daily cases plus the proportions in isolation moved 9 days to the right (b).

**Figure 19:**
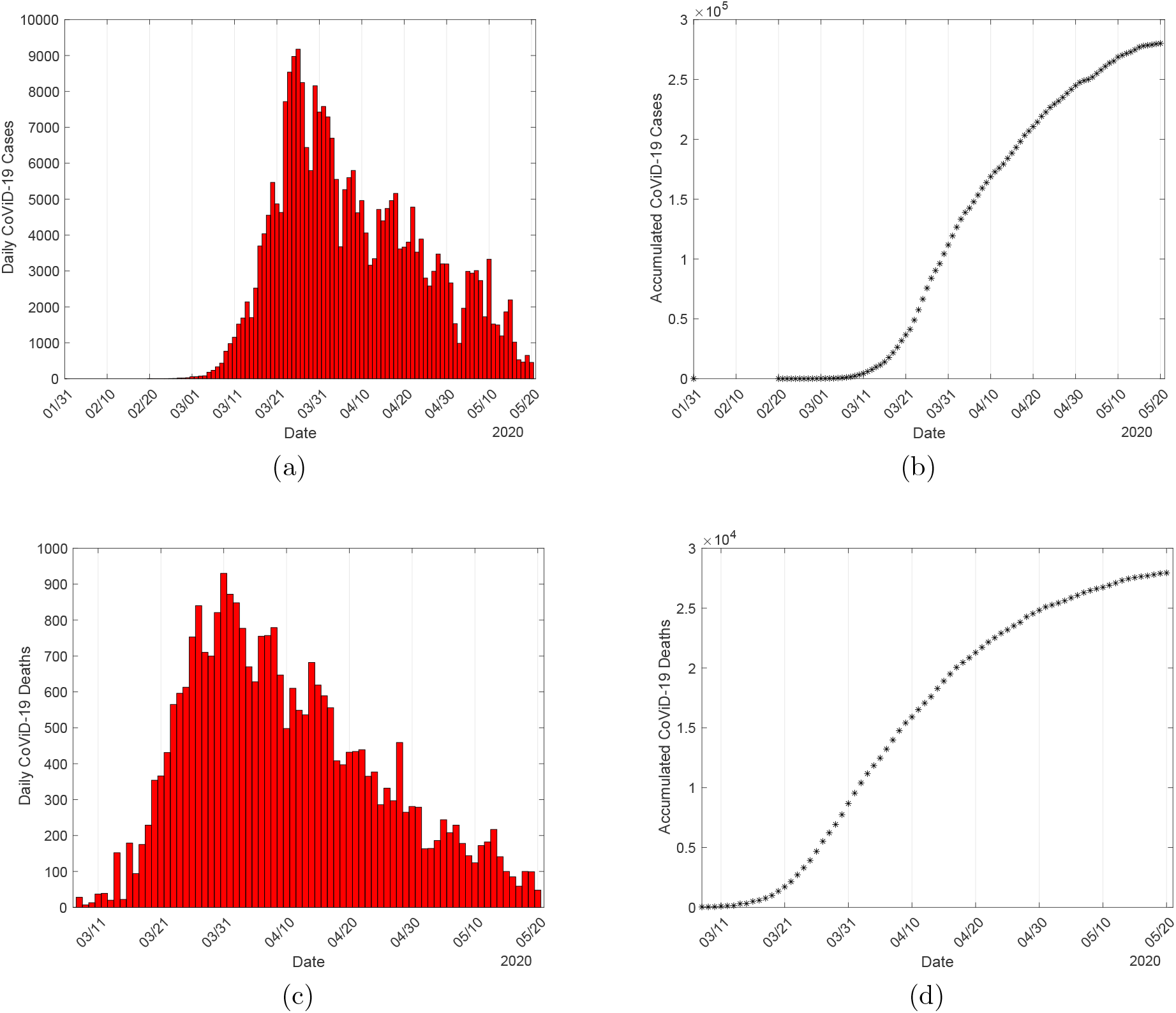
The daily (a) and accumulated (b) covid-19 cases, and daily (c) and accumulated (d) deaths, using covid-19 cases collected in Spain.

The new coronavirus transmission model, based on the above descriptions summarized in Figure 16, is described by the system of ordinary differential equations, with *j* = *y, o*. Equations for susceptible persons are

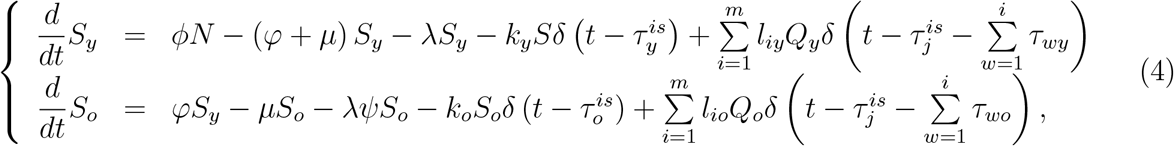

for infectious persons,

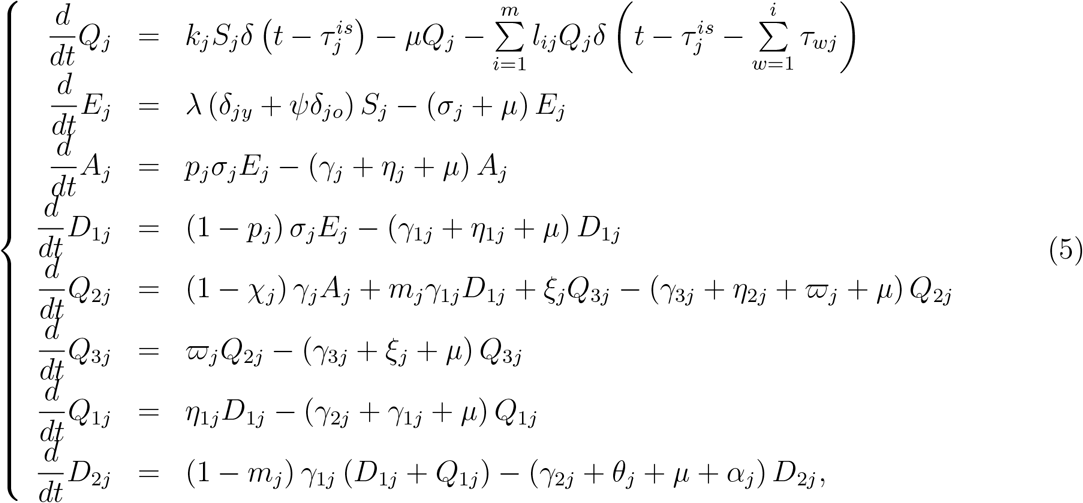

and for immune persons,

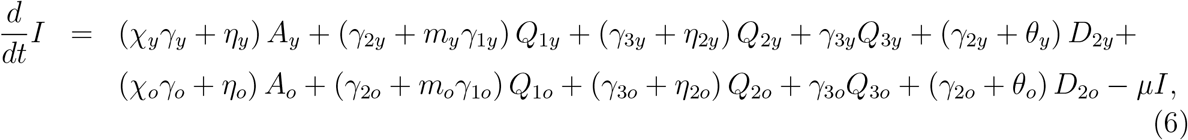

where *N*_*j*_ = *S*_*j*_ + *Q*_*j*_ + *E*_*j*_ + *A*_*j*_ + *D*_*1j*_ + *Q*_*1j*_ + *Q*_*2j*_ + *Q*_*3j*_ + *D*_*2j*_, and *N* = *N*_*y*_ + *N*_*o*_ + *I* obeys

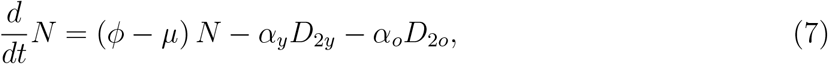

with the initial number of population at *t* = 0 being *N* (0) = *N*_0_ = *N*_0*y*_ + *N*_0*o*_, where *N*_0*y*_ and *N*_0*o*_ are the size of young and elder subpopulations at *t* = 0. If *ϕ* = *µ* + (*α*_*y*_*D*_2*y*_ + *α*_*o*_*D*_2*o*_) */N*, the total size of the population is constant.

Table 1 summarizes the model classes (or variables).

**Table 1:** Summary of the model variables (*j* = *y, o*).

| Symbol | Meaning |
| --- | --- |
| $S_j$ | Susceptible persons |
| $Q_j$ | Isolated among susceptible persons |
| $E_j$ | Exposed and incubating new coronavirus persons |
| $A_j$ | Asymptomatic persons |
| $D_{1j}$ | Pre-diseased (pre-symptomatic) persons |
| $Q_{1j}$ | Pre-diseased persons caught by test |
| $Q_{2j}$ | Mild (non-hospitalized) covid-19 persons |
| $Q_{3j}$ | Mild covid-19 persons adhered to isolation |
| $D_{2j}$ | Severe (hospitalized) covid-19 persons |
| $I$ | Immune (recovered) persons |

**Table 2:**
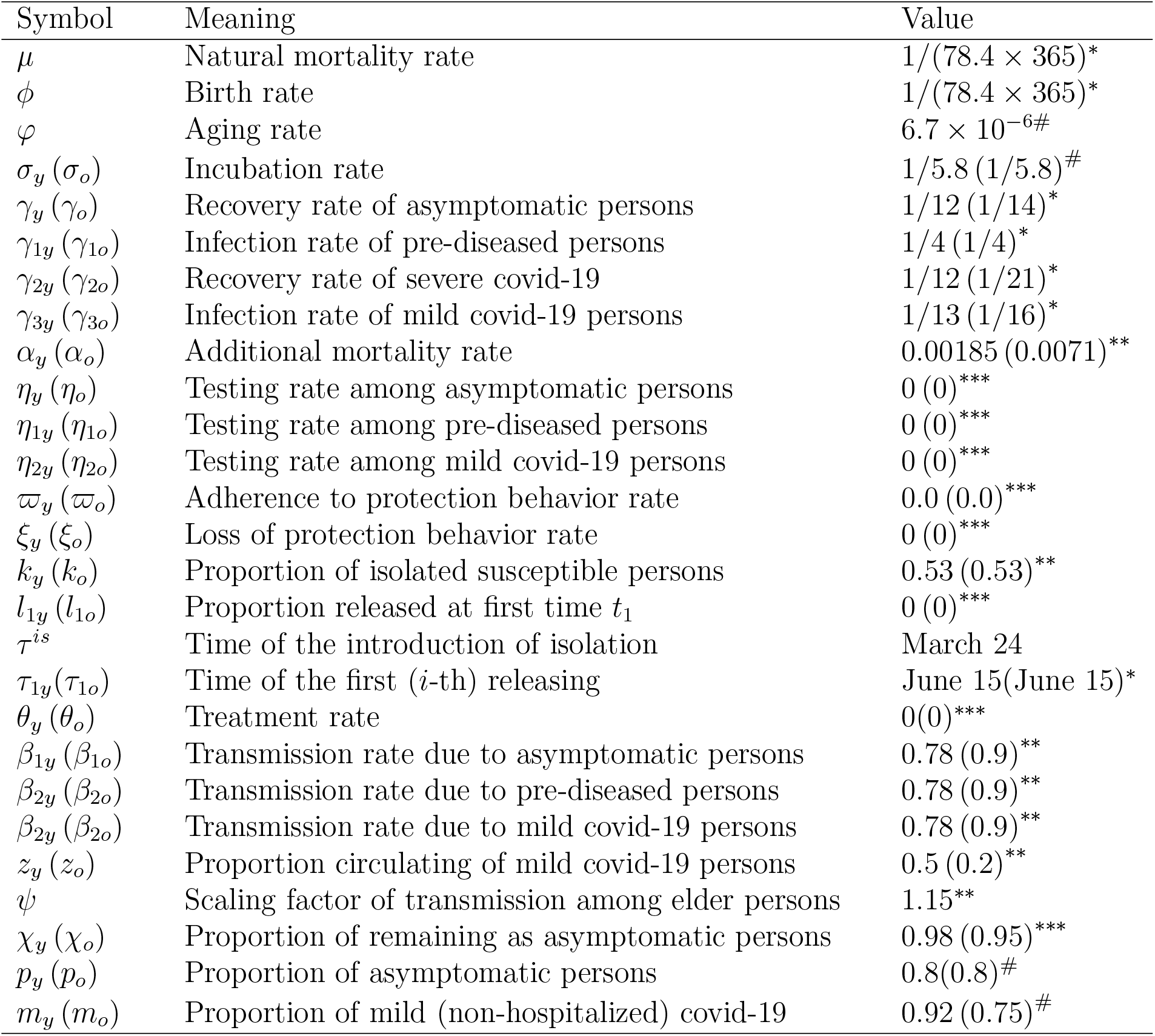
Summary of the model parameters (*j* = *y, o*) and values (rates in *days*^*−*1^, time in *days* and proportions are dimensionless). Some values are calculated (^#^), or obtained from the liteature (^*\**^), or estimated (^*\*\**^) or not available yet (^*\*\*\**^). The life expectancy in Spain is 83.4 *years*.

The system of non-autonomous and non-linear differential equations (4), (5) and (6) is simulated permitting intermittent interventions to the boundary conditions. Hence, the equations for susceptible and isolated persons become

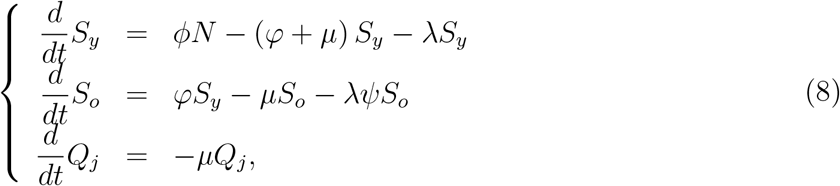

*j* = *y, o*, and other equations are the same. Hence, for the system of equations (5), (6) and (8), the initial conditions (at *t* = 0) are, for *j* = *y, o*,

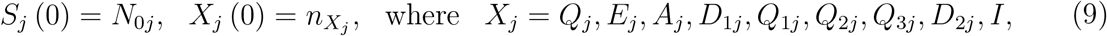

and 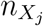 is a non-negative number. For instance, 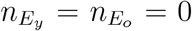means that there is not any exposed person (young and elder) at the beginning of the epidemic.

We do not deal with the release of the isolated population (left to further work), hence here we let *l*_*ij*_ = 0, with *j* = *y, o*, and *i* = 1, 2, *…, m*. The isolation implemented at *t* = *τ* ^*is*^ is described by the boundary conditions

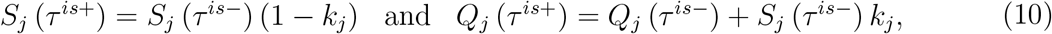

plus

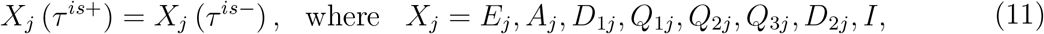

where we have 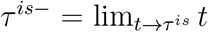, and 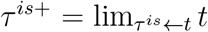. If isolation is applied to a completely susceptible population at *t* = 0, there are not any infectious persons, so *S*(0) = *N*_0_. If isolation is done at 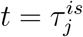 without a screening of persons harboring the virus, then many of the asymptomatic persons could be isolated with susceptible persons.

Table 2 summarizes the model parameters. The description of the assigned values (for elder classes, values are between parentheses) can be found below.

From the solution of the system of equations (5), (6) and (8), we can derive some epidemiological parameters: number of susceptible persons, the number of new cases of covid019, accumulated and daily cases of severe covid-19, and number of deaths due to covid-19. All initial conditions below are determined by the initial conditions (9) supplied to the system of equations.

The number of non-isolated (circulating) persons *S*_*j*_ is obtained from equation (8), and the number of circulating plus isolated susceptible persons *S*^*tot*^ is obtained by

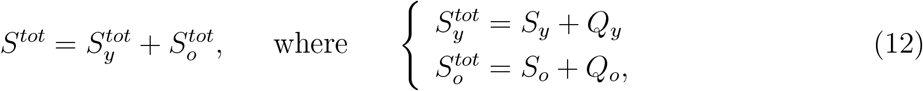

where 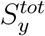 and 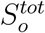 are the numbers of susceptible, respectively, young and elder persons.

The numbers of new cases of covid-19 Φ_*y*_ and Φ_*y*_ are

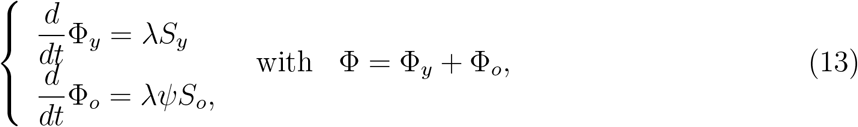

where the force of infection *λ* is given by equation (3).

The numbers of accumulated severe covid-19 cases Ω_*y*_ and Ω_*o*_ are given by the exits from *D*_1*y*_, *Q*_1*y*_, *D*_1*o*_, and *Q*_1*o*_, and entering into classes *D*_2*y*_ and *D*_2*o*_, that is,

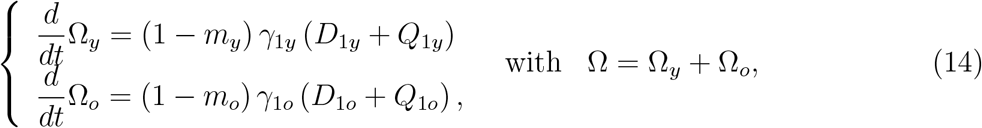

with Ω_*y*_(0) = Ω_*y*0_ and Ω_*o*_(0) = Ω_*o*0_. The daily severe covid-19 cases Ω_*d*_ is, considering Δ*t* = *t*_*i*_ *− t*_*i−*1_ = Δ*t* = 1 *day*,

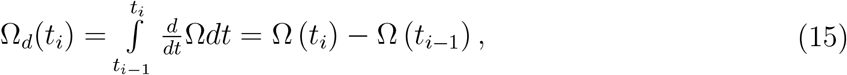

where Ω_*d*_(0) = Ω_*d*0_ is the first observed covid-19 case at *t*_0_ = 0, with *i* = 1, 2, *…*, and *t*_1_ = 1 is the next day in the calendar time, and so on.

The number of deaths due to the severe covid-19 cases is

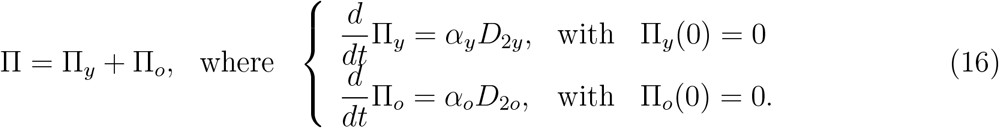

In the estimation of the additional mortality rates, we must bear in mind that the time at which new cases and deaths were registered does not have direct correspondence, rather they are delayed by Δ days, that is, Π_*y*_ (*t* + Δ) = *α*_*y*_*D*_2*y*_(*t*), for instance.

We can estimate the severity case fatality rate as the quotient Π*/*Ω, and the infection fatality rate as Π*/*Φ.

#### The basic reproduction number *R*_0_ – Trivial equilibrium point and its stability

The basic reproduction number *R*_0_ is obtained by the analysis of the trivial equilibrium point in the steady-state. However, the non-autonomous and varying population system of equations (5), (6) and (8) does not have steady-state. Observe that the population in isolation is maintained for a while, hence, disregarding this period of isolation, the system of equations is autonomous (we let *k*_*j*_ = *l*_*ij*_ = 0, *j* = *y, o*), but the population *N* varies. For this reason, considering the fractions of individuals in each compartment defined by

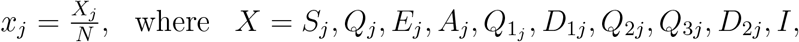

and using equation (7) for *N*, we obtain

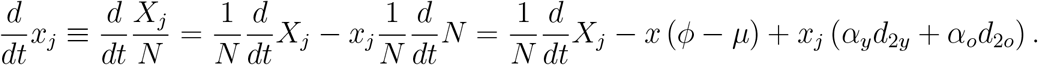

Hence, the system of equations (5), (6) and (8) in terms of fractions become, for susceptible and isolated persons,

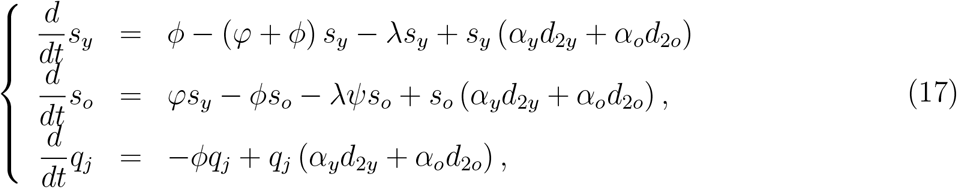

for infected persons,

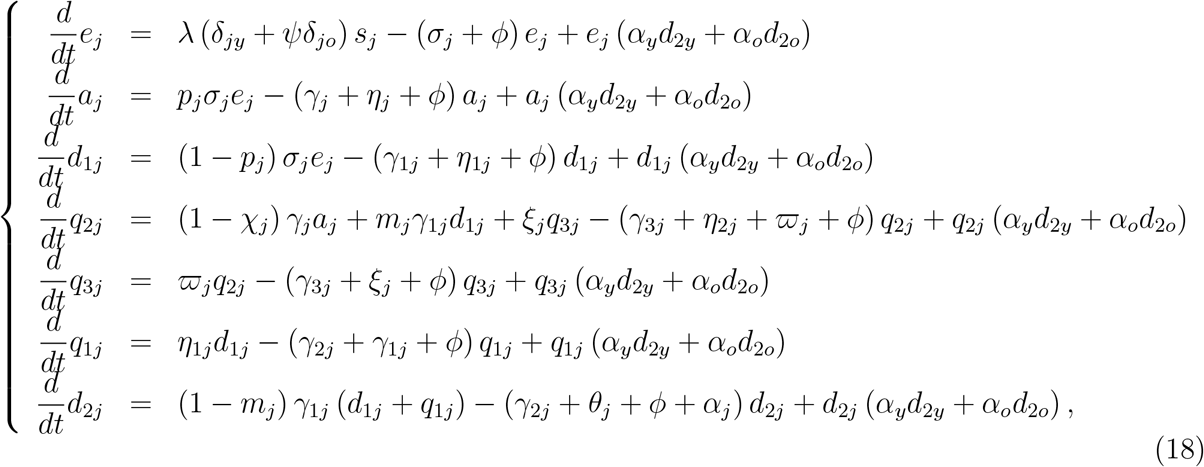

and for immune persons,

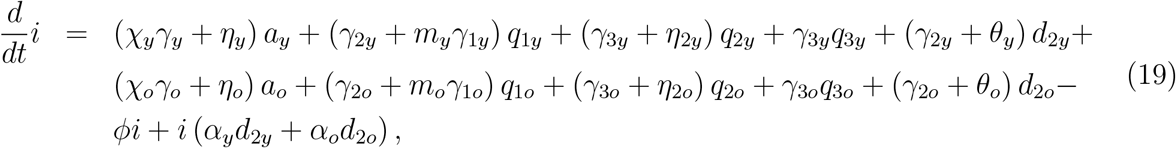

where *λ* is the force of infection given by equation (3) re-written as

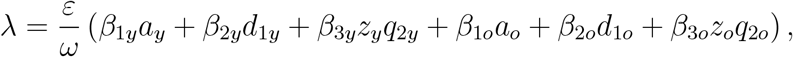

and

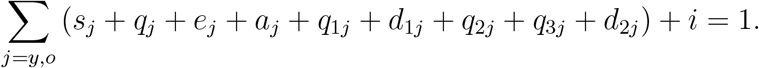

This new system of equation has steady-state, that is, the number of persons in all classes varies with time, however, their fractions attain steady-state (the sum of derivatives of all classes is zero).

The trivial (disease-free) equilibrium point *P* ^0^ of the new system of equations (17), (18) and (19) is given by

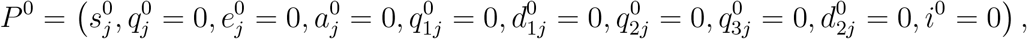

for *j* = *y* and *o*, where

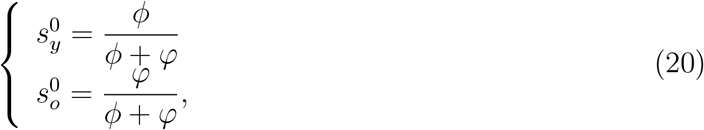

with 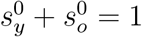.

Let us assess the stability of *P* ^0^ by applying the next generation matrix theory considering the vector of variables *x* = (*e*_*y*_, *a*_*y*_, *d*_1*y*_, *q*_2*y*_, *e*_*o*_, *a*_*o*_, *d*_1*o*_, *q*_2*o*_) [27]. We apply method proposed in [28] and proved in [29]. There are control mechanisms, hence we obtain the reduced reproduction number *R*_*c*_ by interventions.

To obtain the reduced reproduction number, diagonal matrix *V* is considered. Hence, the vectors *f* and *v* are

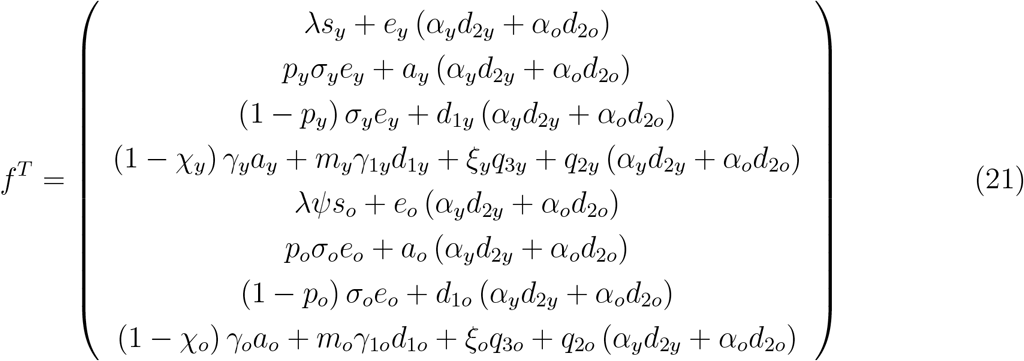

and

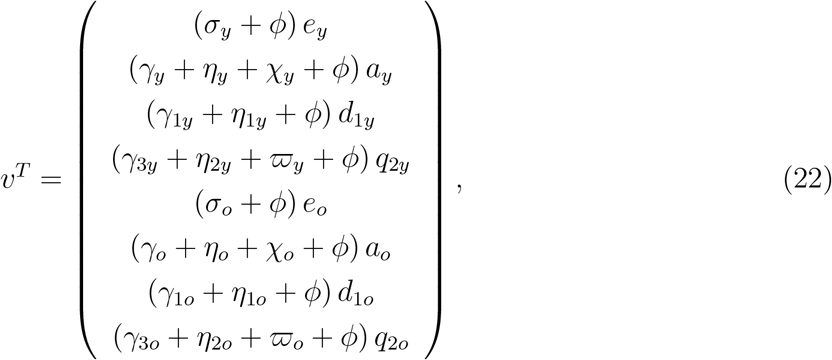

where the superscript *T* stands for the transposition of a matrix, from which we obtain the matrices *F* and *V* (see [27]) evaluated at the trivial equilibrium *P* ^0^, which were omitted. The next generation matrix *FV* ^*−*1^ is

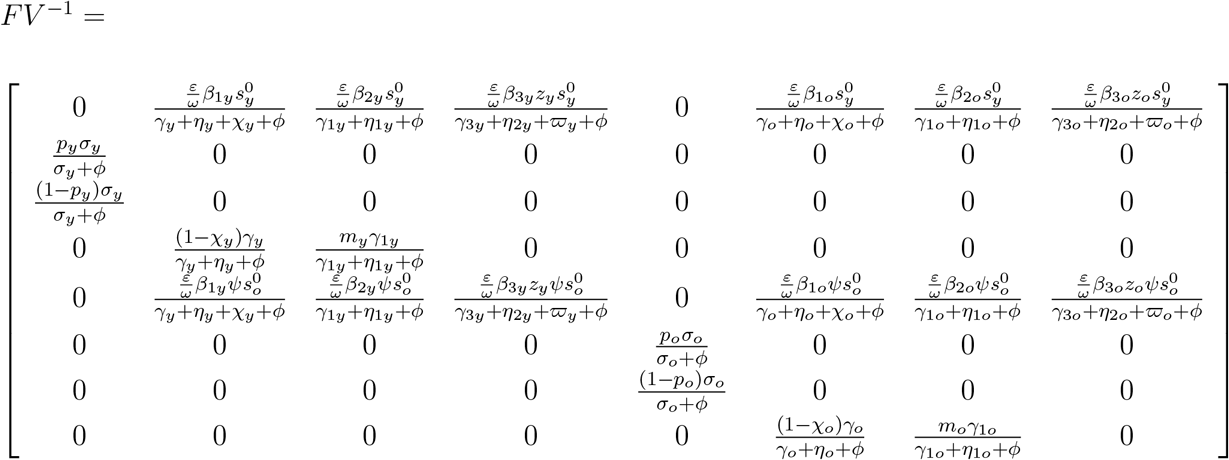

and the characteristic equation corresponding to *FV* ^*−*1^ is

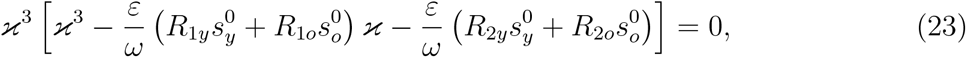

where the partially reduced reproduction numbers *R*_1*y*_, *R*_2*y*_, *R*_1*o*_, and *R*_2*o*_ are

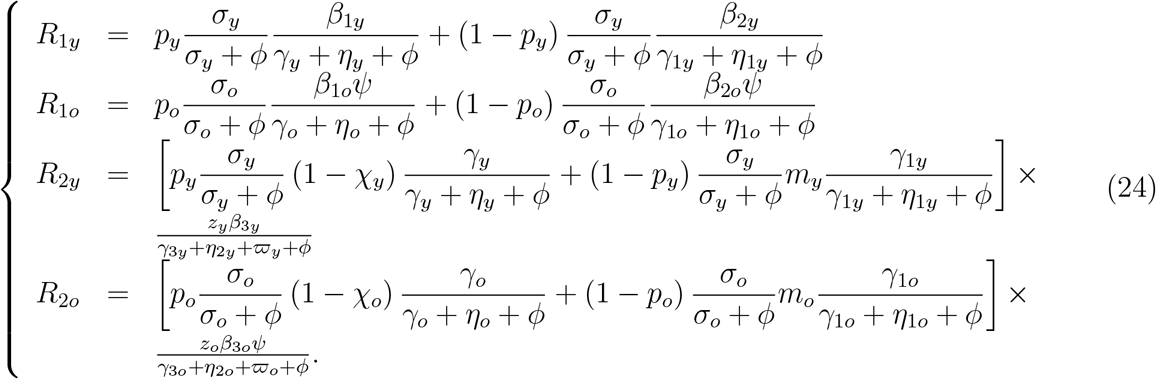

The spectral radius *ρ* (*FV* ^*−*1^) is the biggest solution of a third degree polynomial, which is not easy to evaluate. The procedure proposed in [28] allows us to obtain the threshold *R*_*c*_ as the sum of coefficients of the characteristic equation, where *R*_*c*_ is the reduced reproduction number given by

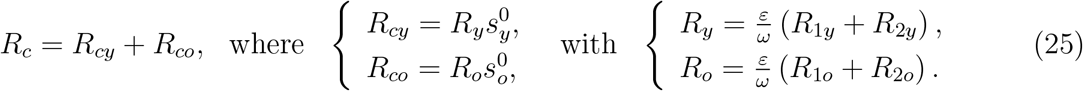

Hence, the trivial equilibrium point *P* ^0^ is locally asymptotically stable if *R*_*c*_ *<* 1.

The basic reproduction number *R*_0_ is given by

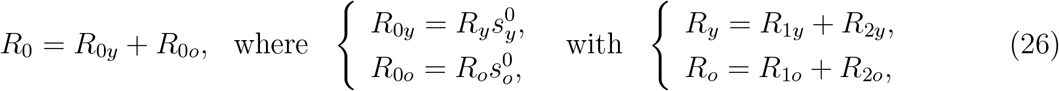

where *R*_1*y*_, *R*_2*y*_, *R*_1*o*_, and *R*_2*o*_ are obtained from equation (24) letting *ε* = *ω* = 1 (absence of protective measures), *η*_*j*_ = *η*_1*j*_ = *η*_2*j*_ = 0 (absence of test), and ϖ_*o*_ = 0 (absence of educational campaign), for *j* = *y, o*. All these control actions to decrease the epidemic are absent at the beginning of the epidemic, and the definition of the basic reproduction number is fulfilled.

The basic reproduction number *R*_0_ is the secondary cases produced by one infectious person (could be anyone in one of the classes harboring virus) in a completely susceptible young and elder populations without constraints [9]. Let us understand *R*_1*j*_ and *R*_2*j*_, *j* = *y, o*, stressing that the interpretation is the same for both subpopulations, hence we drop out subscript *j*. To facilitate the interpretation, we consider this infectious person in exposed class *E*. This person enters into one of the infectious classes composed of asymptomatic (*A*), pre-diseased (*D*_1_), and a fraction of mild covid-19 (*Q*_2_).

1. *R*_1_ takes into account the transmission by one person in asymptomatic *A* or pre-diseased *D*_1_ class. We interpret for asymptomatic person transmitting (between parentheses, for pre-diseased person) infection. One infectious person survives during the incubation period with probability *σ/* (*σ* + *ϕ*) and enters into asymptomatic class with probability *p* (pre-diseased, with 1 *− p*) and generates, during the time 1*/* (*γ* + *ϕ*) (pre-diseased, 1*/* (*γ*_1_ + *ϕ*)) staying in this class, on average *β*_1_*/* (*γ* + *ϕ*) (pre-diseased, *β*_2_*/* (*γ*_1_ + *ϕ*)) secondary cases.
2. *R*_2_ takes into account the transmission by a mild covid-19 person. An infectious person has two routes to reach *Q*_2_: passing through *A* or *D*_1_ (this case is given between parentheses). One infectious person survives during the incubation period with probability *σ/* (*σ* + *ϕ*) and enters into asymptomatic (pre-diseased) class with probability *p* (pre-diseased, with 1 *− p*); survives in this class and also is not caught by a test with probability *γ/* (*γ* + *ϕ*) (pre-diseased, *γ*_1_*/* (*γ*_1_ + *ϕ*)) and enters into mild covid-19 class *Q*_2_ with probability 1 *− χ* (pre-diseased, *m*); and generates, during the time 1*/* (*γ*_3_ + *ϕ*) staying in this class, on average *zβ*_3_*/* (*γ*_3_ + *ϕ*) secondary cases.

Hence, 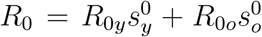 is the overall number of secondary cases generated form one primary case introduced into a completely susceptible young and elder subpopulations. The model parameters are not accurate, and it is expected that *R*_0_ be influenced by the inaccuracy of those values. The variation of *R*_0_ with uncertainties in the parameters can be assessed by the sensitivity analysis [30].

The model given by the system of equations (5), (6) and (8) is non-autonomous and non-constant deterministic model. To obtain the basic reproduction number, we dropped out the pulses in isolation and release in the model and transformed into fractions to obtain *R*_0_. This deterministic model provides an overall view of the epidemic. However, some particular/specific features of the epidemic can be dealt with.

In our model, we can incorporate the mobility especially of infectious individuals, and apply the technique of finding traveling waves. By doing this, the geographic expansion of SARSCoV-2 by calculating the velocity of the front of the wave can be achieved. For instance, the dengue expansion in São Paulo State [31] and West Nile Virus in American Continent [32] were evaluated by the method of finding traveling waves.

We did not consider uncertainties in our model parameters, which can be introduced considering the stochastic version. For instance, the transmission rate *βdt* can be written as 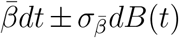, where 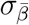 is the standard deviation of average 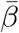 and *B* is the Brownian motion. Gray *et al*. [33] analyzed the SIS model to determine the standard deviation 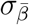 and performed computational simulations.

Instead of continuously varying the number of individuals in each class, our model can be simulated allowing integer variation in the population. For instance, Freitas [34] used a continuous-time stochastic version of the SEIRV (V stands for vaccinated subpopulation) model with integer-valued states. The simulations were done applying the Gillespie algorithm, considering that the transitions between compartments occur randomly according to a Markov chain with transition rates equal to the deterministic rates. A more complex stochastic process can also be applied considering all classes in our model as stochastic variables. For instance, the stochastic process was applied to schistosomiasis transmission [35].

Computational modelings can be formulated. There are innumerous methods, being the agent-based model (ABM) the most widespread. ABM is derived from cellular automata letting each cell carrying on much information. For instance, Ferreira *et al*. [36] applied cellular automata approach to evaluate the suitability of sterile insect technique to control *Aedes aegipty* when breeding sites are homogeneously and heterogeneously distributed.

However, in computational models, after obtaining thousands of trajectories, the mean-field is calculated [36] as well as in the stochastic simulation. However, in the stochastic process, the average values can be calculated for the stochastic variables and, in some cases, these averaged variables follow the corresponding deterministic model [35].

#### The effective reproduction number *R*_*ef*_ – Non-trivial equilibrium point

The effective reproduction number *R*_*ef*_ is defined by one of the equations of the dynamic system at the non-trivial equilibrium point *P*^*\**^. For instance, for the SEIR model, the equation relating susceptible persons must obey at the equilibrium, *R*_0_*s* = 1, from which *R*_*ef*_ is defined by *R*_*ef*_ = *R*_0_*s*, where *s* varies with time. Notice that at the beginning of the epidemic (*t* = 0) we have *s* = 1, and *R*_*ef*_ = *R*_0_, and at the steady-state (*t → ∞*), *s* = *s*^*\**^ and obeys *R*_*ef*_ = 1, resulting in *s*^*\**^ = 1*/R*_0_. In the preceding section we showed that *R*_0_ is given by equation (26), but we did not proved that *s*^*\**^ = 1*/R*_0_ due to the complexity of the new system of equations (17), (18) and (19), and it is not an easy task to determine the non-trivial (endemic) equilibrium point *P*^*\**^.

However, when *z*_*y*_ = *z*_*o*_ = 0, we can show that the inverse of the basic reproduction number *R*_0_ is the fraction of susceptible persons in the steady-state [10]. However, we have young and elder subpopulations, hence the fraction of susceptible individuals at endemic equilibrium 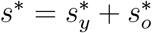 is related generically as

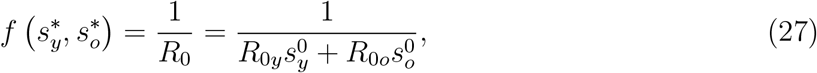

and the effective reproduction number *R*_*ef*_ [37], which varies with time, can not be defined neither by *R*_*ef*_ = *R*_0_ (*s*_*y*_ + *s*_*o*_), nor *R*_*ef*_ = *R*_0*y*_*s*_*y*_ + *R*_0*o*_*s*_*o*_. For instance, for dengue transmission model, 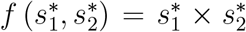, where 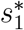 and 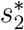 are the fractions at equilibrium of, respectively, humans and mosquitoes [38]. For tuberculosis model considering drug-sensitive and resistant strains, *s*^*\**^ is solution of a second degree polynomial [18]. For this reason, we define the approximated effective reproduction number *R*_*ef*_ as

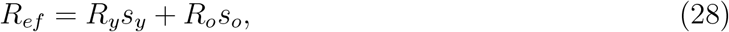

which depends on time, and when attains steady-state (*R*_*ef*_ = 1), we have *s*^*\**^ = 1*/R*_0_.

The basic reproduction number *R*_0_ obtained from mathematical modelings provides two useful information: At the beginning of the epidemic (*t* = 0), *R*_0_ gives the magnitude of control efforts, and when epidemic reaches the steady-state (after many waves of the epidemic, that is, *t → ∞*), *R*_0_ measures its severity providing the fraction of susceptible individuals, in general, *s*^*\**^ = 1*/R*_0_ [18]. Between these two extremes, the effective reproduction number *R*_*ef*_ dictates the course of an epidemic, which follows decaying oscillations around *R*_*ef*_ = 1 [14].

The effective reproduction number *R*_*ef*_, given by equation (28), can be applied directly to the system of equations (5), (6) and (8) by substituting *s*_*y*_ and *s*_*o*_ by *S*_*y*_*/N* and *S*_*o*_*/N*, that is,

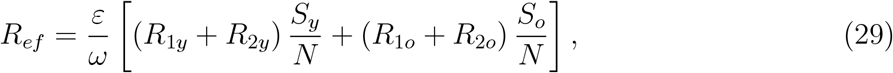

where *S*_*y*_, *S*_*o*_, and *N* vary with time (remember that the fractions *S*_*y*_*/N* and *S*_*o*_*/N* approach steady-state). We describe the variation of *R*_*ef*_ during the epidemic.

1. Natural epidemic. The population, initially, did not adopt any protection measures against the covid-19 epidemic, and at *t* = 0 we have *R*_*ef*_ (0) = *R*_0_, where the basic reproduction number is obtained by substituting 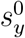 and 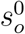 by *N*_0*y*_*/N*_0_ and *N*_0*o*_*/N*_0_ in equation (20), resulting in

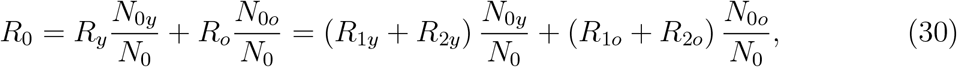

where *R*_*y*_ and *R*_*o*_ are given by equation (26). For *t >* 0, *R*_*ef*_ decreases as susceptible populations decrease.
2. Isolation. At *t* = *τ* ^*is*^ a pulse in isolation is introduced, decreasing the number of susceptible persons, from *S*_*y*_ (*τ* ^*is−*^) and *S*_*o*_ (*τ* ^*is−*^) to *S*_*y*_ (*τ* ^*is*+^) and *S*_*o*_ (*τ* ^*is*+^), see equations (10) and (11). The decrease in the susceptible populations at *τ* ^*is*^ results in *R*_*ef*_ (*τ* ^*is−*^) jumping down to *R*_*ef*_ (*τ* ^*is*+^) = *R*_*r*_, where the reduced reproduction number *R*_*r*_ is given by

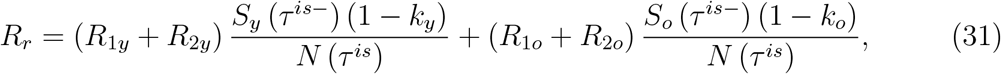

where *R*_1*y*_, *R*_2*y*_, *R*_1*o*_, and *R*_2*o*_ are given by equation (26). For *t > τ* ^*is*^, *R*_*ef*_ decreases as susceptible populations decrease.
3. Adopting protective measures. The protective measures are incorporated in the modeling by the factor *ε* in the circulating population and the restricted contact by *ω* in the isolated population. At the time of introducing these measures *T*, the effective reproduction number just before the time of the adoption of protective measures *R*_*ef*_ (*T*^*−*^) jumps down to *R*_*ef*_ (*T* ^+^) = *R*_*p*_ by factor *ε/ω*, that is,

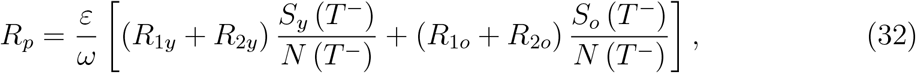

where *R*_1*y*_, *R*_2*y*_, *R*_1*o*_, and *R*_2*o*_ are given by equation (26). For *t > T, R*_*ef*_ decreases as susceptible populations decrease.

Therefore, the importance of a mathematical model is the capability of providing the basic and effective reproduction numbers, which give information about the risk of infection during the epidemic. Our model is complex, but we could obtain *R*_0_, which was not an easy task. However, a more complex model can be formulated incorporating novel findings of the covid-19 epidemic, but we must be aware that the basic reproduction number must be obtained from the model.

### Description of data collection from São Paulo State and Spain

We first present a detailed description of the data collected from São Paulo State, and briefly the data collected from Spain.

#### Data from São Paulo State

Figure 17 shows the daily (a) and accumulated (b) severe covid-19 cases, and daily (c) and accumulated (d) deaths collected from São Paulo State [17].

Figure 18 shows the proportion in the population in isolation (a), and daily cases plus the proportion in isolation moved 9 days to the right (b). The horizontal line in Figure 18(a) corresponds to the mean value *k*_*mean*_ = 0.53, around which daily proportions vary impacting on the transmission [39].

We discuss roughly the collected covid-19 cases shown in Figure 17. Interestingly, Figure 17(a) shows that the daily data present weekly seasonality, with lower cases at the weekend [17], due to the procedure of registering the day at which occurred the confirmation by laboratory testing, not the beginning of symptoms.

A. The number of SARS in São Paulo State registered in the site of Ministry of Health (Brazil) [40] shows increasing beyond the average cases occurred in past years since March 8, 2020 (around 1, 000 cases in the 11^*th*^ epidemiological week (hereafter, week), March 8-14), and reach peak 2 weeks later (around 4, 000 cases in the 13^*th*^ week, March 22-28). After this epidemiological week, the notification as SARS initiates decreasing trend, maybe due to increased testing of severe covid-19 cases (on March 31, there were 822 cases, but one day earlier, only 66 cases, and around 180 cases per day in the 13^*th*^ week). Figure 17(b) shows this jump up on March 31. This increased number of cases should be explained by more testing among SARS to identify covid-19, or by the exponential-like increasing of the epidemic in the beginning, or probably by both. Figure 17(a) shows an unusual jump up when comparing 13^*th*^ week and 14^*th*^ week (March 29 - April 4), which is not observed in the next weeks, suggesting that the isolation decreased the force of infection. Indeed, the isolation was introduced on March 24, but after 10 days, on April 3, there is a change in the exponential-like trend, becoming less abrupt. Figure 17(b) shows an increasing trend in blocks of week affected by weekly seasonality shown in Figure 17(a) depending on the proportion in isolation that occurred 9 or 10 days earlier.
B. Let us compare severe covid-19 cases and proportion in isolation week by week, Figure 17(b). Notice that there is a jump up from the 13^*th*^ to 14^*th*^ week, showing an exponential-like increase. However, there is not jump in the 15^*th*^ week (April 5-11), possibly showing the effects of isolation. In the 16^*th*^ week (April 12-18) there were strong variations in the covid-19 cases, maybe due to huge variation in the proportions in isolation 9 days earlier. In the 17^*th*^ week (April 19-25), the increased number of cases corresponds to decreased proportions in isolation including weekend (on April 25, Sunday, there was the highest number of cases). This increasing trend continued in the next 18^*th*^ week (April 26 - May 2), when the proportions in isolation fluctuated, but a relatively small number of cases were registered during the extended holiday (May 1-3). The behavior observed in the 17^*th*^ and 18^*th*^ weeks may be the effects of manifestation against isolation occurred on April 18: the peak on April 25 and the high number of cases lasting until April 30, that is, 7 to 12 days after the manifestation is the interval with median 9.5 and variation 2.5 (sum of incubation and pre-diseased periods is 9.8).
C. From Figure 17(b), the accumulated covid-19 cases show three periods with different trends. The first period from February 26 to April 3 corresponds to the natural epidemic. The second period, from April 4 to April 12, corresponds to the isolation effectively decreasing the epidemic, and the last period, since April 13, the additional reduction in the transmission of covid-19 occurs due to the protective measures.

From the daily registered deaths due to covid-19 shown in Figure 17, we observe a correspondence between the number of deaths, Figure 17(c), and the number of covid-19 cases, Figure 17(a), occurred around 15 days ago. For instance, on May 18, São Paulo State registered 41 deaths, and on May 19, 324. However, on May 3 and 4, 15 and 14 days before May 18, there were registered, respectively, 598 and 415 covid-19 cases, while on May 5 and 6, 14 and 13 days before May 19, 1, 866 and 3, 800 covid-19 cases were registered. Hence, we can infer that the number of deaths corresponds to cases of covid-19 occurred around 14 to 15 days earlier. For instance, on May 13, 14, 15, and 16, the daily registered covid-19 cases were, respectively, 3, 378, 3, 189, 4, 092, and 2, 805, and it is expected an elevated number of deaths after 15 days.

#### Data from Spain

Figure 19 shows the daily (a) and accumulated (b) severe covid-19 cases, and daily (c) and accumulated (d) deaths collected from Spain, from January 31 to May 20 [21].

Analyzing the data collected from Spain, we observe three trends. The first period, from January 31 to March 21, had an exponential-like increase, which is followed by the second period with less increase from March 22 to 28, and the last period with a slow increase since March 29. The effects of lockdown implemented on March 16 are expected to appear 9 days later [15], on March 25. The intermediate period from March 22 to 28 is centered on March 25 with 3 days of variation, called the transition period from natural to lockdown epidemic. Hence, we divide the epidemiological scenarios of Spain into three stages, and we estimate the transmission rates *β*_*y*_ and *β*_*o*_, the protection factor *ε*, and the decreasing factor *ω*. The additional mortality rates *α*_*y*_ and *α*_*o*_ are also estimated.

### Values for the model parameters

The values assigned in Table 2 are obtained from the estimation of covid-19 cases and deaths, or they are calculated.

#### Model parameters evaluated from the data

We model parameters can be fitted applying the least square method (see [41]), that is,

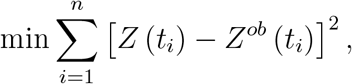

where min stands for the minimum value, *n* is the number of observations, *t*_*i*_ is *i*-th observation time, *Z* stands for Ω given by equation (14), or Π given by equation (16); and *Z*^*ob*^ stands for the observed number of severe covid-19 cases Ω^*ob*^ or the number of died persons Π^*ob*^. However, this estimation method of parameters is extremely complex, and considering observed data only in one variable of the dynamic system is not appropriate (see [42]). For this reason, we evaluate the sum of squared differences by varying model parameters. This simplified method of parameter evaluation does not provide uncertainties associated with the parameters.

To evaluate the transmission rates in the natural epidemic (there are not any interventions at the beginning of the epidemic, then *ε* = *ω* = 1), we assume that all rates in young persons are equal, as well as in elder persons letting

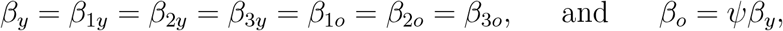

and the forces of infection are *λ*_*y*_ = (*A*_*y*_ + *D*_1*y*_ + *z*_*y*_*Q*_2*y*_ + *A*_*o*_ + *D*_1*o*_ + *z*_*o*_*Q*_2*o*_) *β*_*y*_*/N* and *λ*_*o*_ = *ψλ*. The reason to include a factor *ψ* is the reduced capacity of a defense mechanism by elder persons (physical barrier, innate and adaptive immune responses, etc.). Another fact is the expression of angiotensin-converting enzyme 2 in the nasal epithelium, the receptor that SARS-CoV-2 uses for host entry, which increases with age [43]. The force of infection takes into account all virus released by infectious individuals (*A*_*y*_, *D*_1*y*_, *Q*_2*y*_, *A*_*o*_, *D*_1*o*_ and *Q*_2*o*_), the rate of encounter with susceptible persons (many factors, especially demographic density and closeness), and the capacity to infect them (see [44] [45]). Additionally, the amount inhaled by susceptible persons can be determinant in the chance of infection and the prognosis of covid-19 [46]. We also assume that the proportions in the isolated young and elder subpopulations are equal, that is, *k* = *k*_*y*_ = *k*_*o*_.

Currently, there is not a sufficient number of kits to detect infection by the new coronavirus. For this reason, test to confirm infection by this virus is done only in hospitalized persons, and also in persons who died manifesting symptoms of covid-19. Hence, we have only data of accumulated severe covid-19 cases (Ω = Ω_*y*_ + Ω_*o*_) and those who died (Π = Π_*y*_ + Π_*o*_).

To evaluate the parameters *β*_*y*_ and *β*_*o*_ = *ψβ*_*y*_, the proportions in the isolated population *k*, and reduction in the transmission rates due to protective measures adopted by population *ε* and decreased contact *ω*, we calculate

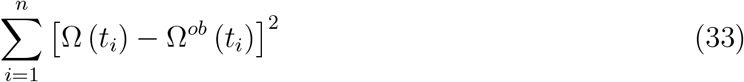

by varying the parameters, and we choose the lower sum of the squared distances between the curve and data. The accumulated covid-19 cases Ω is given by equation (14).

To estimate the mortality rates *α*_*y*_ and *α*_*o*_, the let *α*_*y*_ = Γ*α*_*o*_, where Γ is provided by the ratio of deaths occurring in young and elder subpopulations. We fix the previously estimated parameters *β*_*y*_ and *β*_*o*_, *k, ε*, and *ω*, and evaluate

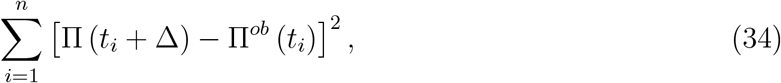

by varying the parameter *α*_*o*_. The accumulated deaths due to covid-19 Π is given by equation (16), with Π(0) = 0. Notice that the time of registration *t*_*i*_ of deaths must be related to the deaths of new cases Δ times ago, that is, *D*_2_(*t*_*i*_*−*Δ). We use Δ = 15 *days* obtained by analyzing the data from São Paulo State.

#### Assignment of values for the model parameters

Some values for model parameters are found in the literature, and other parameters are calculated.

For incubation period, we use mean value between 5.2 [47] and 6.4 [48], that is, *σ* = *σ*_*y*_ = *σ*_*o*_ = 1*/*5.8 *days*^*−*1^. We use for the infectious rates of pre-diseased persons, *γ*_1_ = *γ*_1*y*_ = *γ*_1*o*_ = 1*/*4 *days*^*−*1^ [49], which is indirectly confirmed by the delay observed in 9 days between low isolation and increase in covid-19 cases. It was observed approximately 2 weeks for the duration of mild disease, then we use *γ*_*o*_ = 1*/*14 and *γ*_*y*_ = 1*/*12 (both in *days*^*−*1^), and critical disease lasts 2-6 weeks, then we use *γ*_2*o*_ = 1*/*21 and *γ*_2*y*_ = 1*/*12 (both in *days*^*−*1^) [3].

Li *et al*. [8] observed that 86% of all infections were undocumented, and assuming that the ratio between asymptomatic and symptomatic young and elder persons are equal, we let *p* = *p*_*y*_ = *p*_*o*_ = 4*/*5 = 0.8. Additionally, Pan *et al*. [50] observed that in a family of three, one had clinical symptoms, and the other two members were both asymptomatic. From São Paulo State, 76% of deaths due to covid-19 are 60 years old or more, then the ratio of death is 1 : 3 to young persons [17]. However, the ratio may be lower in severe covid-19 cases, then we assume 2 : 3 (in Sao Bernardo de Campo City, São Paulo State, the ratio of hospitalized young and elder persons is 2 : 3.3). From the observation that 81% are mild and can recover at home [51], we consider a lowered ratio between mild and severe covid-19 among elder persons 3 : 1, hence *m*_*o*_ = 3*/*4 = 0.75. To have ratio 2 : 3 between young and elder in hospitalized persons, we must have approximately 10 : 1 in the ratio between asymptomatic and symptomatic among young persons. Gathering the above information, we calculate approximately

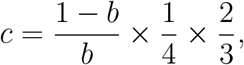

where the term (1 *− b*) */b* is the populational ratio between young and elder persons, 1*/*4 is the proportion of severe covid-19 cases among elder persons, and 2*/*3 is the ratio between hospitalized young and elder persons. Using *b* = 0.153, we have *c* = 0.92. Hence, the ratio asymptomatic:symptomatic of young persons is approximately 12 : 1, which results in *m*_*y*_ = 12*/*13 = 0.92, for *p*_*y*_ = *p*_*o*_.

The dynamic of the new coronavirus propagation is obtained by evaluating the system of equations (5), (6) and (8) numerically using the 4^*th*^ order Runge-Kutta method. Let us determine the initial conditions supplied to this system. In São Paulo State, the number of inhabitants is *N* (0) = *N*_0_ = 44.6 million [11]. The value of parameter *φ* given in Table 1 was calculated by rewriting the equation (20) as *φ* = *bϕ/* (1 *− b*), where *b* is the proportion of elder persons. Using *b* = 0.153 in São Paulo State [11], we obtained *φ* = 6.7 *×* 10^*−*6^ *days*^*−*1^, hence, *N*_*y*_ (0) = *N*_0*y*_ = 37.8 million 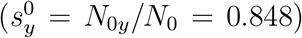 and *N*_*o*_ (0) = *N*_0*o*_ = 6.8 million 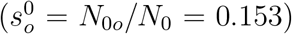. Hence, the initial conditions for susceptible persons are *S*_*y*_ (0) = *N*_0*y*_ and *S*_*o*_ (0) = *N*_0*o*_ for São Paulo State. Following the same idea, the initial conditions for Spain are *S*_*y*_ (0) = 35.17 million and *S*_*o*_ (0) = 12.23 million.

The initial conditions for other variables are calculated based on Table 2. Using *p*_*y*_ = *p*_*o*_ = 0.8, the ratio asymptomatic:symptomatic is 4 : 1 for young and elder persons; using *m*_*o*_ = 0.75, the ratio mild:severe covid-19 is 3 : 1 for elder persons, and for young persons, the ratio is 12 : 1 from *m*_*y*_ = 0.92. Hence, for elder subpopulation, if we assume that there is one person in *D*_2*o*_ (the first confirmed case), then there are 3 persons in *Q*_2*o*_; the sum (4) is the number of persons in *D*_1*o*_, implying that there are 16 in *A*_*o*_, hence, the sum (20) is the number of persons in *E*_*o*_. Notice that, if there is 1 person in *D*_2*y*_, then there must be 12 persons in *Q*_2*y*_. For young subpopulation, we assume that there is not any person in *D*_2*y*_, but 6 persons in *Q*_2*y*_, then the sum (6) is the number of persons in *D*_1*y*_, implying that there are 24 in *A*_*y*_, hence, the sum (30) is the number of persons in *E*_*y*_. Finally, we suppose that no one is isolated or tested, and immunized.

Therefore, the initial conditions supplied to the dynamic system (5), (6) and (8) are, for young and elder subpopulations,

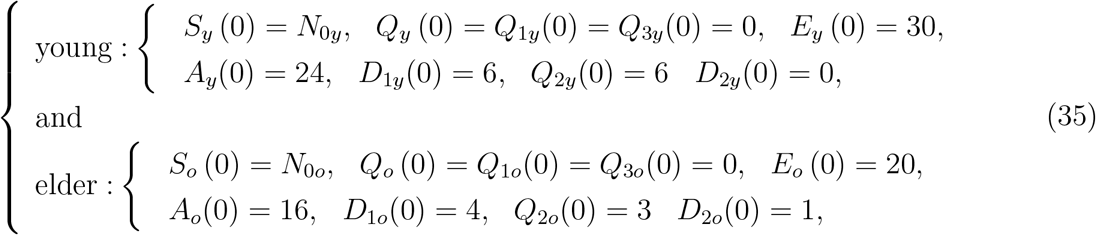

plus *I*(0) = 0, where the initial simulation time *t* = 0 corresponds to the calendar time when the first case was confirmed (February 26 for São Paulo State, and January 31 for Spain).

## Data Availability

Data collected from Såo Paulo State government and Spain Health Ministry

https://www.saopaulo.sp.gov.br/coronavirus/

https://www.mscbs.gob.es/profesionales/saludPublica/ccayes/alertasActual/nCov-China/situacionActual.htm

## Notes

### Competing Interest Statement

The authors have declared no competing interest.

### Funding Statement

None

